# Putting Polygenic Scores in Context: How Intersectional Factors Affect Relative and Absolute Genetic Risk

**DOI:** 10.1101/2025.06.04.25328987

**Authors:** Mihael Cudic, Justin D. Tubbs, Tian Ge, Jordan W. Smoller

## Abstract

**Background:** The clinical utility of polygenic scores (PGS) is known to vary when training and test samples differ in ancestry, with recent work suggesting that sociodemographic differences can also impact PGS performance. However, the impact of belonging to multiple intersecting contexts on genetic risk remains understudied.

**Methods:** We analyzed lifetime disease odds ratios (OR) and absolute risks (AR) in high-PGS individuals across 106 two-way intersections of sociodemographic factors (sex, age, alcohol intake, smoking, income, deprivation). Seven diseases were assessed: atrial fibrillation, coronary artery disease, type 2 diabetes, hypercholesterolemia, asthma, obesity, and major depression. Primary analyses were performed in the *UK Biobank* (n=375,054, British-European-like ancestry), with replication in *All of Us* (n=36,552, African-like; n=99,477, European-like).

**Results:** ORs varied significantly across contexts, with greater variation observed across intersectional contexts. On average, the maximal OR variation across two-way contexts was 56%. AR deviations were more moderate after adjusting for context prevalence but still showed intersectional effects. For example, high-PGS, low-income individuals had an average 1.0 percentage-point drop in estimated AR across phenotypes using a context-aware vs. context-unaware PGS, while those additionally reporting low alcohol intake had a 3.1-point lower AR estimate for major depression in UKB. Results were generally consistent across datasets, with strongest replication in European-like AoU samples.

**Discussion:** Our findings show that intersectional contexts can have a sizable impact on genetic risk effect estimates. Future clinical applications may need to incorporate these contextual effects to improve accuracy and fairness for patient-specific genetic risk assessment.

## Introduction

Polygenic scores (PGS) are powerful tools for capturing individual-level variation in common genetic variants and estimating an individual’s genetic risk for a specific trait or condition ^1^. Over the past decade, numerous studies have established that PGS are robust risk factors for a broad range of health conditions, leading to growing interest in the potential clinical application of PGS for informing risk stratification and prevention ^2–6^. For example, the eMERGE consortium has developed and implemented a pipeline for incorporating PGS into risk assessment for multiple diseases with return of results to patients and clinicians ^6^. A widely-known challenge facing clinical application of PGS is their limited portability across populations that differ in ancestry ^7–9^, with the predictive power of PGS generally declining as the genetic distance between the target and training samples increases ^1,10^. This limitation is particularly important because differences in PGS prediction accuracy across populations may affect the clinical utility of chosen cutoffs, potentially leading to disparities in risk classification and medical decision-making.

Although ancestry is a well-recognized contributor to disparities in PGS accuracy, it is only one instance of a broader phenomenon in which mismatch between training and target samples due to contextual factors can influence predictive performance. Recent studies have shown that ancestry, sex, age, and socioeconomic status (SES) can all affect PGS accuracy at both individual ^11,12^ and population levels ^13,14^. For example, Mostafavi et al ^14^ conducted a series of analyses showing that PGS performed differently across contexts within ancestry groups: PGS for diastolic blood pressure were less predictive in males than in females; PGS for body mass index performed worse in older individuals than in younger ones; and PGS for years of schooling were less predictive in higher socioeconomic status (SES) contexts compared to lower SES contexts. Another recent hypothesis-free study targeted at PGS prediction of BMI found 18 environmental contexts where prediction accuracy differed significantly ^13^. Recently, Hou et al. ^11^ proposed a method to adjust individual-level prediction intervals around PGS to reflect differences in accuracy across contexts. However, these studies have considered only the linear effects of each context and have not modeled the potential effects of interactions between contexts on PGS. Furthermore, they focus solely on prediction accuracy without examining how these disparities translate into clinically-relevant absolute risks, leaving a gap in understanding the real-world implications of context-aware PGS risk estimations.

In this work, we address these limitations through a series of analyses to explore the impact of sociodemographic contexts on relative and absolute genetic risk. First, we examine how relative genetic risk varies across differing contexts among individuals with high PGS, as defined by the eMERGE consortium ^6^. Specifically, we pay particular attention to intersectional contexts, recognizing that individuals exist within multiple overlapping dimensions, to better quantify the variability of genetic risk estimates for individuals of diverse backgrounds. Second, we evaluate how well these intersectional contextual effects generalize across biobanks (UK Biobank vs. All of Us) and ancestries (European-like, African-like). Finally, we evaluate more clinically-relevant performance of a context-aware PGS by analyzing their impact on the estimated absolute risk of high-PGS individuals. Overall, this study offers key insights into how intersections of contextual factors might influence the interpretation and application of genetic risk in healthcare.

## Methods

### Phenotypes and Polygenic Scores

Phenotypic and genetic data for our primary analysis were collected through the UK Biobank (UKB) ^15^, a population-based sample of approximately 500,000 individuals with comprehensive multi-modal data including surveys, interviews, linked electronic health records, and array-based genotyping. Data used in this work were obtained under UKB application #32568.

For the current study, we restricted our analyses to a set of unrelated white British ancestry participants while also adjusting for genetic principal components (PCs) to minimize potential effects of population stratification. Individuals and variants flagged by the UKB’s genetic quality control pipeline were excluded prior to analysis. Up to 374,024 individuals of white British ancestry with high-quality imputed genetic data were available for analysis.

Seven disease phenotypes were examined: atrial fibrillation (AFIB), coronary artery disease (CAD), type 2 diabetes (T2D), hypercholesterolemia (HC), asthma (AST), obesity (OB), and major depressive disorder (MDD). Cases were identified through a combination of ICD codes recorded in linked hospital records (Fields 41202 or 41204) and self-reported disease history (Field 20002). Supplemental Table 1a lists the ICD and self-report codes used to identify cases for each disease or disorder, along with the sample sizes for each phenotype, which ranged from 355,187 to 374,024. PGS weights were calculated by applying PRS-CS-auto ^16,17^ to summary statistics from the largest available GWAS (not containing UK Biobank participants) for each of the seven traits (Supplemental Table 3). Subsequently, mean-centered and variance-standardized PGS were calculated using PLINK version 1.9 ^18^.

For replication analyses, we used data from the *All of Us* Research Program (AoU) ^19^, an ongoing longitudinal study for which multi-modal data were available on over 240,000 individuals, including surveys, physical measurements, linked electronic health records, and whole genome sequencing data. We restricted these replication analyses to individuals of European-like or African-like ancestry, the two largest genetic ancestry groups available in AoU. After filtering variants and individuals flagged for removal by AoU researchers, we calculated PGS using the same PRS-CS weights as described above, mean-centering and standardizing the PGS within each ancestry group separately. Two sets of within-sample genetic PCs were also calculated using PLINK version 1.9 within European and African ancestry samples separately. Cases for each phenotype were identified as those with the presence of at least one diagnostic code for the disease or disorder in the linked electronic health records, as defined by the SNOMED codes shown in Supplemental Table 1b. Additionally, to align the AoU cohort with the UK Biobank cohort, we restricted the AoU sample to individuals aged 40 and older, matching UKB’s minimum age. The available sample sizes for each of the 7 conditions available in AoU among African and European ancestry participants ranged from 36,552 to 99,477, and are listed in Supplemental Table 1b.

### Intersectional Contexts

Six primary sociodemographic variables available in the UKB were used to contextualize PGS: sex, age, alcohol intake, smoking history, household income, and Townsend deprivation index (Supplemental Table 2). Context strata were categorized as low, medium, and high levels within these seven primary environmental variables, except for binary variables of sex and lifetime smoking history. We attempted to create strata which resulted in relatively evenly-distributed sample size while maintaining meaningful definitions. For example, the Townsend deprivation index was split into contexts of low (<−3; n=141,325), middle (−3 to 0; n=133,498), and high (>0; n=99,784) strata. For smoking history, the two categories consisted of never smokers (n=201,738) and previous or current smokers (n=171,745) to balance sample sizes across strata.

In total, we considered 16 primary sociodemographic context strata. From these, 106 two-way intersectional contexts were derived through pairwise combinations of each primary context and were used as the main unit of analysis throughout this study. Three-way intersectional contexts were also included if they contained more than 50 cases to ensure sufficient sample size. A detailed breakdown of the sample sizes for each intersectional stratum is provided in Supplemental Table 1a.

The AoU replication analysis included the same phenotypes, as well as both one-way and two-way intersectional contexts (Supplemental Table 1a). Similar to the UKB analyses, we categorized continuous and ordinal variables into low, medium, and high strata, aiming to balance participant distribution across the three categories while maintaining interpretability. The mapping between UKB and AoU variables, along with the corresponding participant counts, is provided in Supplemental Table 2.

### Estimating Genetic Risk

The lifetime relative and absolute risk of disease attributable to PGS within a given context was assessed using both the odds ratio (OR) and absolute risk (AR) for individuals with the highest PGS, as proposed by the PGS cutoffs defined by eMERGE ^6^ (Supplemental Table 3). Since no recommended PGS cutoff exists for MDD, we used the top 1% of PGS as the high genetic risk cutoff.

Computing OR and AR using high PGS cutoffs within stratified groups poses risks of positivity violations and overfitting due to small sample sizes within strata. These challenges are especially pronounced in AoU, where the smaller cohort sizes further increase the likelihood of these issues arising. To address this, we applied an approximation method ^20^ that estimates OR and AR using only the variance explained (R²) by PGS on the observed scale and disease prevalence.

This approach has several advantages over the more standard logistic regression method. First, this method allows for the estimation of genetic risk at the extreme ends of the distribution, under the assumption that the PGS follows a normal distribution. Second, because OR and AR are derived from the PGS R², we are able to further mitigate overfitting within strata by regressing out covariates across the entire sample rather than within each stratum separately. Finally, this approach is more computationally efficient, making it feasible to perform extensive bootstrapping to more accurately estimate statistical significance. We compare the OR estimates between the Pain et al. method ^20^ and logistic regression in Supplemental Figure 1. Overall, the OR estimates from both methods show strong correspondence, but this agreement weakens when examining smaller subsets of the top percentiles of the PGS. Notably, substantial deviations in OR estimates appear when case counts are low within a given stratum, which may suggest that either logistic regression is more prone to overfitting in such scenarios, or that PGS tails deviate from the normality assumption.

Therefore, genetic risk was computed within each stratum as follows. Each PGS and phenotype were first residualized by regressing out the top 10 principal components (PCs), genotyping chip (if available), and each of the six demographic and environmental context variables in the full sample. Participants were then stratified based on their measured sociodemographic and environmental variables. For this residualization step, missing values were mean-imputed. However, imputed variables were not used for stratum assignment or any subsequent analyses. We then performed linear regressions using the residualized PGS and phenotype for participants within each stratum to calculate the PGS R² on the observed scale after accounting for linear effects of PCs and demographic and environmental variables. The observed R^2^, along with the prevalence of the condition in that stratum, was finally used to estimate OR and AR at the recommended PGS cutoff off points using the method of Pain et al. (2022) ^20^.

Because clinicians and patients may be more interested in absolute as opposed to relative risks of disease, we compared three estimates of AR for high-PGS individuals within a specific context *c*. The first estimate, *AR*_*unaware*_, represents the risk for high-PGS individuals without accounting for their specific context, as traditionally done in PGS studies. This estimate is based on the overall relative disease risk (*OR*_*overall*_), as derived from Pain et al. (2022), and the disease prevalence *prev*_*overall*_ estimated in the full sample, and is calculated as follows:

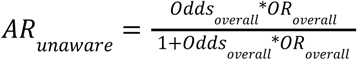

where *Odds*_*overall*_ is defined as:

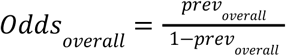

The second AR estimate, *AR*_*adj*_, accounts for the prevalence of disease within the specific context (*prev_c_*), but assumes that the relative genetic risk remains constant across contexts. As a result, it does not incorporate the impact of contextual factors on PGS. It is calculated as follows:

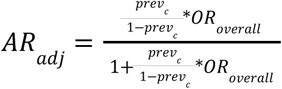

Finally, the third estimate, *AR*_*c*_, is derived from a context-aware PGS by adjusting for the effect of contextual factors on PGS. This estimate uses the relative genetic risk within that context (*OR_c_*), as again derived from Pain et al. (2022), and is computed as:

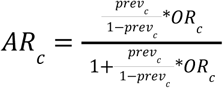

Since *AR*_*c*_ and *AR*_*adj*_ are calculated using the same disease prevalence but differ in how they account for relative genetic risk, the differences between these two estimates (*AR*_*c*_ - *AR*_*adj*_) represents the change in absolute risk for high-PGS individuals when their PGS effect is contextualized.

### Assessing the Variation of ORs and the Importance of Variables

To determine the extent of the variability in ORs across different contexts, we computed the percent difference in ORs between the stratum with the highest and lowest OR among a given set of context strata. This measure, referred to as the maximum OR %Δ, was defined as:

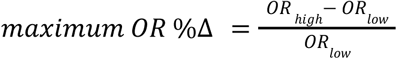

The maximum OR %Δ was used for two different types of analysis. First, it was used to assess variability in ORs across one-way, two-way, and three-way intersections of contextual variables. Specifically, we calculated the percent difference between the highest and lowest OR within each set of contextual strata: all 16 one-way contexts, all 106 two-way intersectional contexts, and three-way contexts with at least 20 cases. This last analysis was conducted only in UKB due to the insufficient sample size in AoU for analyzing three-way intersectional contexts.

Second, the maximum OR %Δ was used to measure the impact of a specific variable or variable interactions on relative genetic risk. For example, for alcohol intake, the maximum OR %Δ was calculated across all 3 alcohol-related contexts: low intake, medium intake, and high intake while for the interaction of alcohol intake and income, the maximum OR %Δ was calculated across all 9 alcohol-by-income intersectional contexts. In total, we calculated the maximum OR %Δ for all 6 individual variables and all 15 two-way variable intersections. For each cohort (UKB Eur., AoU Afr., and AoU Eur.), the highest and lowest OR stratum were specific to that cohort and did not necessarily align across cohorts.

### Determining Statistical Significance

We assessed the significance of the maximum OR %Δ by empirically calculating p-values and confidence intervals using 10,000 bootstrapped samples. To generate these samples, we repeatedly sampled participants within the predefined minimum and maximum OR strata, which were identified from the full-sample analysis. Therefore, the minimum and maximum strata remained fixed throughout bootstrapping.

To evaluate the significance of intersectionality on relative genetic risk, we directly compared the maximum OR %Δ bootstrapped distributions across all one-way, two-way and three-way contexts with one another. Statistical significance was assessed using Bonferroni correction to account for the three pairwise comparisons per phenotype: p<.05/3 *, p<.01/3 **, p<.001/3 ***.

For assessing the significance of maximum OR %Δ across all 6 variable and 15 two-way variable intersections, bootstrapped distributions were compared to a null value of 0, representing no difference in OR across the set of strata. Bonferroni corrections were applied to account for the 21 (6 variable and 15 two-way intersection) comparisons, with significance thresholds set at p<.05/21 *, p<.01/21 **, p<.001/21 ***.

To determine whether interactions between two variables significantly influenced PGS performance beyond simple additive effects, we compared the bootstrapped distributions of maximum OR %Δ values from their two-way intersections to the full-sample point estimate maximum OR %Δ of each parent variable. For example, we directly compared the max. OR %Δ across all 9 alcohol-by-income intersectional contexts with the max. OR %Δ across all 3 alcohol-related contexts and again with all 3 income-related contexts. Therefore, two empirical p-values—one for each parent variable– were calculated using the bootstrapped samples described above. We employed a one-tailed test, assessing whether the two-way interaction effect was greater than the effects of both parent variables. If the maximum of these two p-values, i.e., max(p₁, p₂), was < .05, the interaction was considered significant (denoted as ⨂ in Figure 3). This additional test was only performed on two-way variable intersections that had already been deemed significant themselves, as described in the previous paragraph, to minimize multiple comparisons. The max. OR %Δ for the two-way intersection and the parent variables were bootstrapped separately, such that there was no correspondence between distributions.

Statistical significance of the Spearman’s Rank correlations of ORs between cohorts were determined by bootstrapping the correlations across all 106 intersectional contexts. To eliminate potential biases arising from differences in PGS predictive power for a given phenotype, the mean two-way intersectional OR for each phenotype was subtracted from that phenotype’s ORs across all contexts for each cohort. Given the three pairwise cohort comparisons (UKB European vs. AoU European, UKB European vs. AoU African, and AoU European vs. AoU African), Bonferroni correction was applied to adjust for multiple testing. Significance thresholds were denoted as follows: * p<.05/3, ** p<.01/3, *** p<.001/3.

Finally, to identify and evaluate significant deviations in AR estimates, we selected the top 5 largest positive and negative differences between *AR*_*c*_ and *AR*_*adj*_ within a given two-way context for each phenotype within UKB. From these, we then identified the top 8 phenotype-context pairs with the largest AR deviations observed in UKB that were also at least nominally significant (p<.1) in both UKB and AoU European-like ancestry cohorts. Statistical significance was then assessed using a Bonferroni correction accounting for 6 total comparisons: two types of AR differences (*AR*_*c*_-*AR*_*unaware*_ and *AR*_*c*_-*AR*_*adj*_) evaluated across the three cohorts. As a result, the corrected significance thresholds were set to * p < .05/6, ** p < .01/6, and *** p < .001/6.

## Results

Our analysis reveals substantial variation in estimated genetic risk of high-PGS individuals across sociodemographic intersectional contexts, highlighting the importance of incorporating intersectionality in genetic risk calibration. For illustration, we use the example of AFIB which has a recommended high-risk threshold at or above the top 3% of the PGS distribution, as defined by the eMERGE consortium. In UKB, individuals with a PGS in the top 3% have an OR of 3.31 (Figure 1). To achieve the same clinically actionable OR in those aged 40–49, the cutoff would need to be redefined as the top 6.26% of PGS values. As more contextual factors are considered, further adjustments are needed. For men aged 40–49, the high-risk cutoff expands to include the top 10.99% of PGS. Among men aged 40-49 with no smoking history, the cutoff yet again expands to the top 15.11% of PGS. This highlights the importance of considering intersectional contexts when determining high genetic risk thresholds to avoid inappropriately excluding individuals who actually meet the intended level of risk.

**Figure 1:**
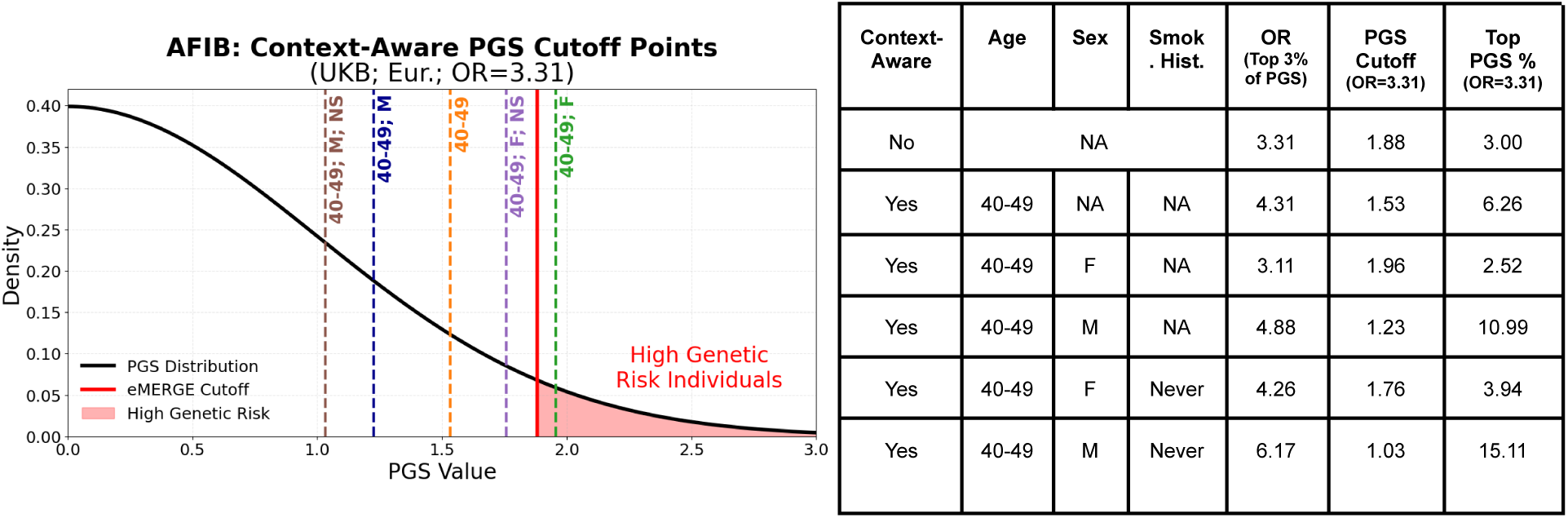
Context-aware PGS cutoff points for atrial fibrillation in the UK Biobank. **a)** A normal distribution of polygenic score (PGS) values for atrial fibrillation (AFIB) in the UK Biobank (UKB) is shown in black. The solid red line represents the eMERGE-recommended PGS cutoff (top 3%) for identifying individuals at high genetic risk. Dashed lines represent context-aware PGS cutoffs for individuals aged 40–49, further stratified by sex (females [F] and males [M]) and smoking status (never smoked [NS]). These context-aware PGS cutoffs were calibrated so that each subgroup maintained the same level of relative genetic risk as the overall population cutoff used in eMERGE, corresponding to an odds ratio (OR) of 3.31. ORs were calculated without adjustment for covariates to maintain consistency with the original eMERGE cutoff definition, which was based on unadjusted relative genetic risk estimates. **b)** A table presenting the ORs for individuals in different subgroups classified as high genetic risk based on the eMERGE top 3% cutoff for AFIB. The table also includes the context-aware PGS cutoffs necessary to achieve an OR of 3.31.

The full span of ORs for AFIB in UKB high-PGS (top 3%) individuals across one-way and two-way intersectional contexts is depicted in Figure 2a. ORs span from 2.94 to 3.61 in one-way contexts and broaden to 2.22 to 4.01 in two-way intersectional contexts. As illustrated in Figure 2b, this range expands further in three-way intersectional contexts, while maintaining statistical significance. The pattern of increasing OR variability with greater intersectionality holds across all phenotypes analyzed in the UKB and is statistically significant for most (Figure 2c). On average, the maximum OR %Δ nearly doubles with each additional layer of intersectionality: 26.2% [95% CI: 21.6%–31.2%] across one-way contexts, increasing to 56.0% [95% CI: 44.0%–71.4%] across two-way intersections, and more than doubling again to 141.7% [95% CI: 104.0%–203.0%] across three-way intersections.

**Figure 2:**
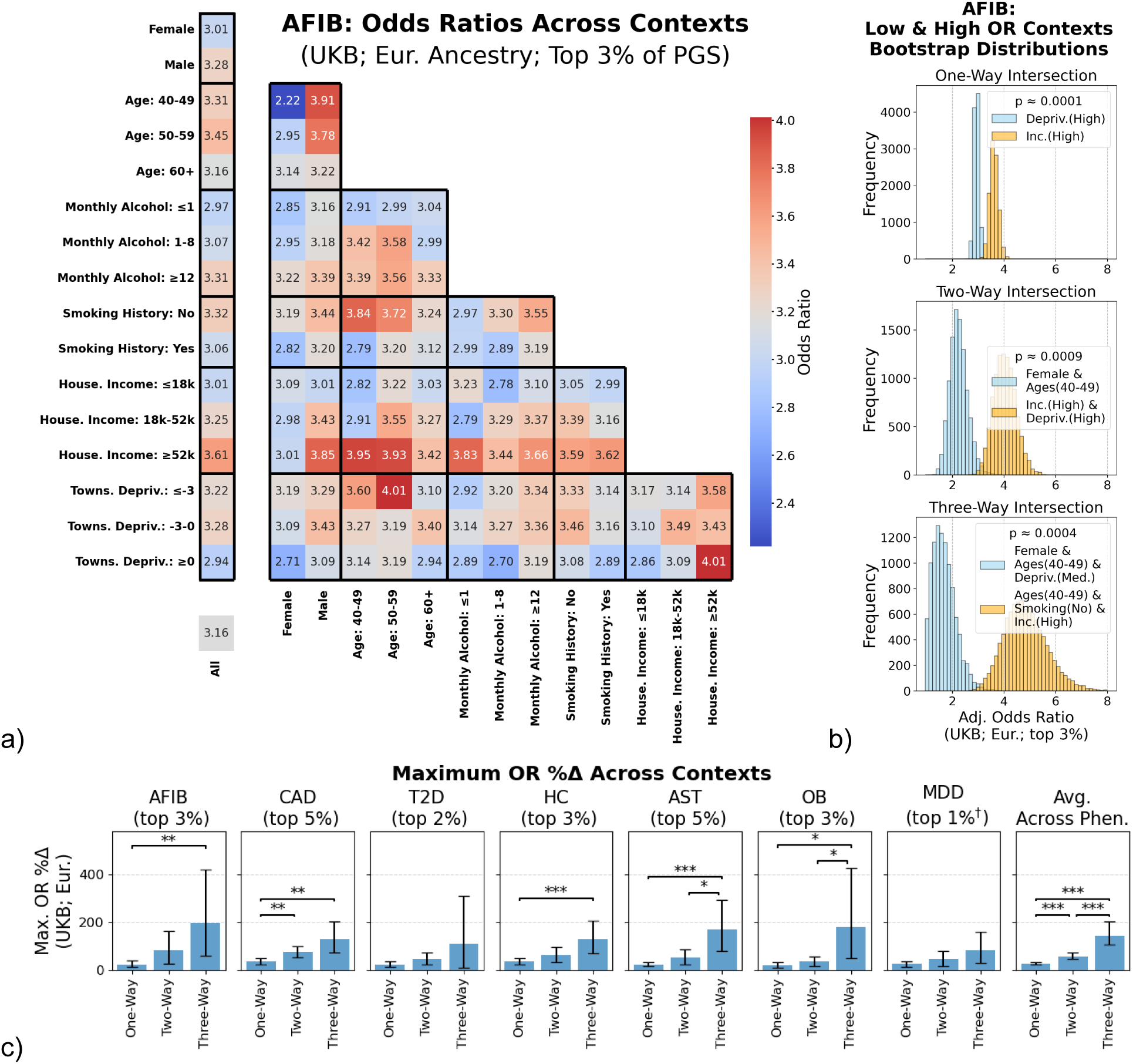
Relative genetic risk differs substantially across contexts and increases in variability as more contextual factors are intersected. **a)** The ORs for individuals in the top 3% of PGS overall, as determined by the recommended eMERGE PGS cutoff, across varying one-way and two-way contexts for AFIB within UKB European ancestry. **b)** Bootstrapped distributions (n=10,000) of the ORs from the contexts with the lowest and highest ORs across one-way, two-way, and three-way analyses. **c)** The percent differences between the lowest and highest OR contexts (max. OR %Δ) across one-way, two-way, and three-way intersectional contexts for all phenotypes using the eMERGE PGS cutoffs († MDD PGS cutoff was set to 1% as a recommendation was not made). Statistical significance between max. OR %Δ for a given phenotype was empirically determined from the bootstrapped distributions (n=10,000; * p<.05/3, ** p<.01/3, *** p<.001/3).

When analyzing the effect of individual variables and their two-way intersections on relative genetic risk, we found that variable intersections can both have a significant effect themselves and a significantly greater effect than either parent variable alone. For instance, in the case of AFIB among UKB individuals, the maximum OR %Δ across all sex-related and age-related strata was 9.1% [95% CI: 1.7%, 17.1%] and 9.2% [95% CI: 0.4%, 18.7%], respectively, neither of which was significant (Figure 3a). However, when considering all six sex-by-age strata together, the maximum OR %Δ reached 76.2% [95% CI: 24.9%, 150.9%], which was both statistically significant (p≈9e-4) and significantly greater than the effect of either sex or age alone on relative genetic risk (p≈4.2e-3). Figure 3b further illustrates this effect, showing that alcohol intake and smoking history significantly influence relative risk, but mainly when interacting with other variables. A comprehensive visualization of ORs and ARs across different contexts and the maximum OR %Δ across variables for all remaining phenotypes and cohorts—including the AoU European and African Ancestry cohorts—is provided in Supplemental Figures 2-8. Additionally, Supplemental Table S1 (`Supplemental_Table_S1.xlsx`) provides detailed OR estimates for all phenotypes and cohorts, while Supplemental Table S2 (`Supplemental_Table_S2.xlsx`) summarizes the maximum OR %Δ across variables.

**Figure 3:**
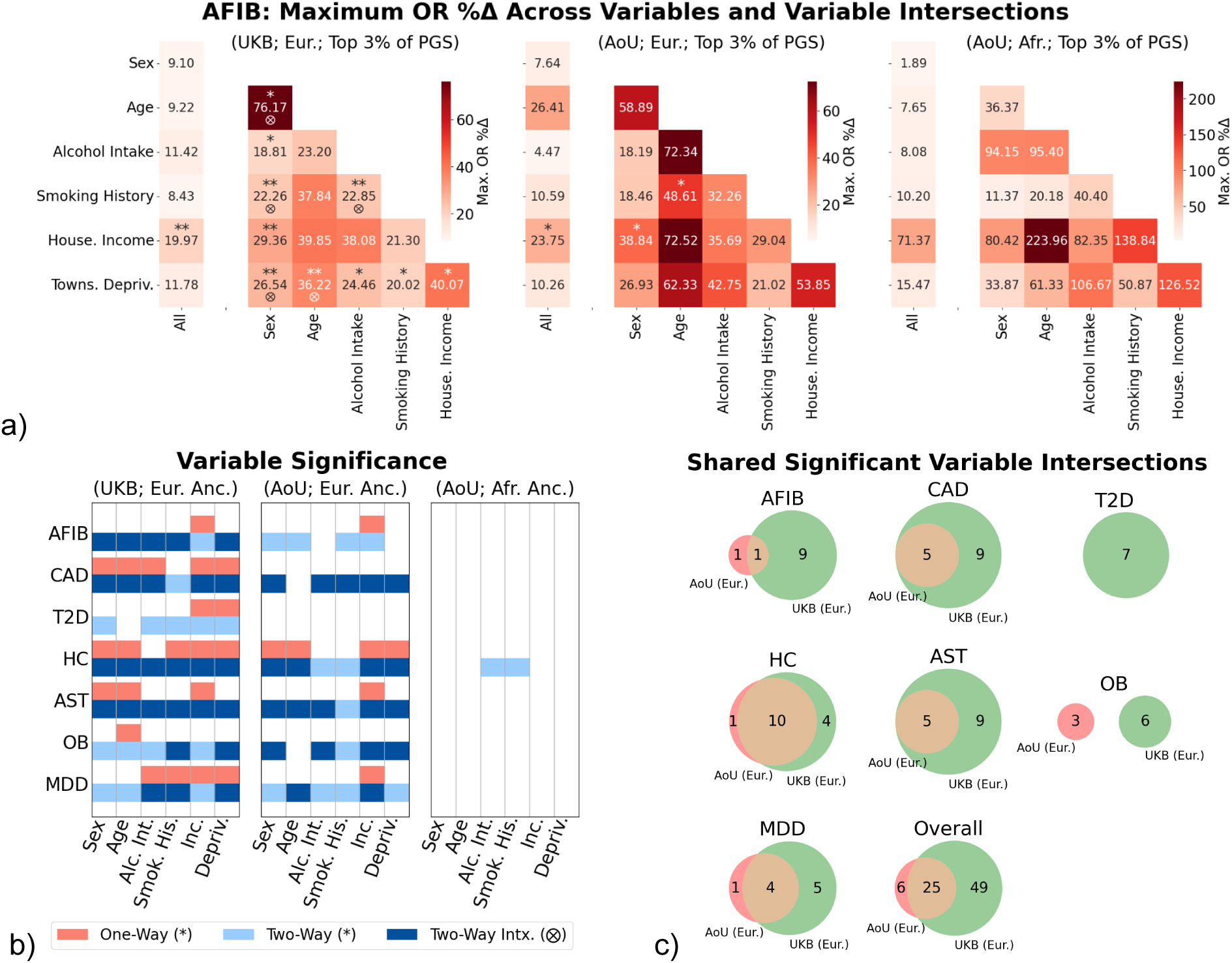
Significant variable and variable intersections were largely consistent across biobanks, especially within ancestry. **a)** The maximum percent difference in odds ratio (max. OR %Δ) across strata, calculated either within a single variable (i.e., across all strata associated with that variable) or at the intersection of two variables (i.e., across all strata formed by combinations of two variables), for AFIB in three cohorts: UKB European ancestry (*left*), AoU European ancestry (*middle*), and AoU African ancestry (*right*). The statistical significance of a variable or the intersection of two variables was empirically determined using a bootstrapped distribution of the max. OR %Δ (n=10,000; * p<.05/21, ** p<.01/21, *** p<.001/21). Additionally, an interaction between two variables was considered significant if the max. OR %Δ for their intersection was statistically significant both on its own (i.e., * p<.05/21) and when compared to the max. OR %Δ of each individual variable (n=10,000; ⨂ p < .05). **b)** Variable significance, determined by *p*-values from max. OR %Δ bootstrapped distributions are shown for all phenotypes across the three cohorts. Variables are categorized as follows: independently significant (* p<.05/21; salmon), significant in combination with another variable (* p<.05/21; light blue), and significant due to an interaction effect with another variable (⨂ p < .05; dark blue). **c)** The overlap between significant intersections of two variables (where max. OR %Δ p < .05/21) between UKB and AoU European ancestry for all phenotypes and overall. Note, the overlap with AoU African ancestry intersections was not shown as only one intersection (HC; alcohol intake and smoking history) was determined to be significant.

The AoU replication results for African and European ancestry show substantial similarity between the UKB and AoU European findings, as illustrated in Figure 3. Similar to the UKB analysis, certain PGS × context variables—such as alcohol intake and smoking history—that were not individually significant became significant in intersectional analyses within the AoU European cohort. Additionally, as shown in Figure 3c, 25 out of 31 two-way intersections with significant maximum OR %Δ in the AoU European cohort were also significant in the UKB European cohort. In contrast, the AoU African ancestry cohort yielded only one significant two-way intersection, likely due to lower statistical power.

Additionally, there were significant correlations in the ORs across biobanks and ancestries for all 106 two-way intersectional contexts (Figure 4). After adjusting for differences in phenotype-specific ORs, the strongest overall correlation was observed within the same ancestry (European) between AoU and UKB biobanks (r=.41; [95% CI: .35, .47]) (Figure 4a and 4b). In contrast, the weakest overall correlation, though still significant, was found across both ancestry and biobanks (UKB European vs. AoU African ancestry; r=.13; [95% CI: .05, .20]) (Figure 4b). An example of this strong correlation can be seen in Figure 4c, where CAD exhibits a consistent pattern across income and sex, with the OR increasing with income for males but not for females. However, ORs were not always aligned, as seen with AST across strata combining alcohol intake and age (Figure 4d). Although the maximum OR %Δ across all nine alcohol-by-age strata was significant in both European UKB and AoU cohorts for AST, the ORs within individual strata differed considerably between the two cohorts. Distributions of the OR correlations between cohorts for strata within each of the 15 two-way variable intersections are illustrated in Supplemental Figure 9.

**Figure 4:**
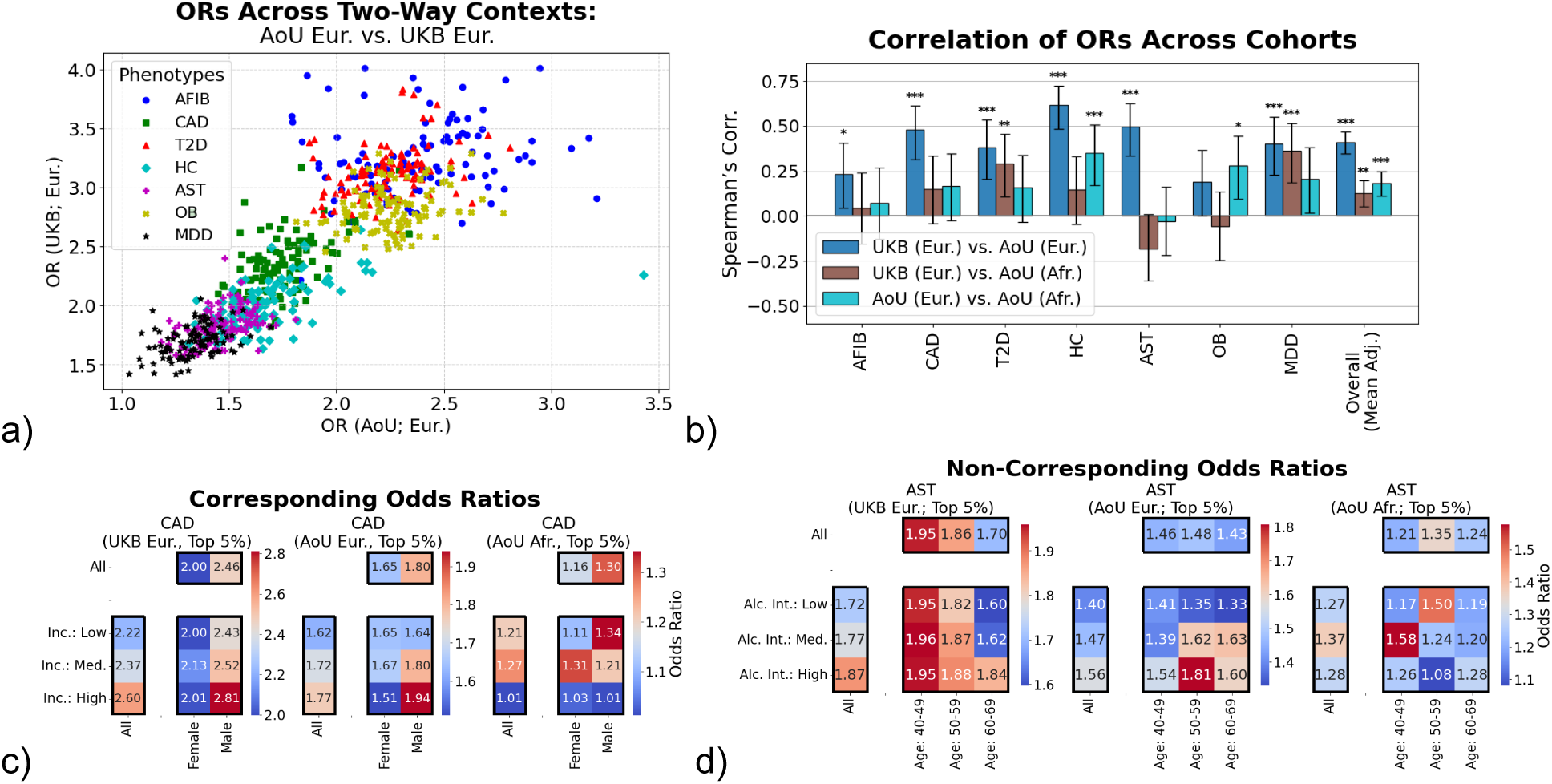
Correspondence and non-correspondence of relative genetic risk across intersectional contexts between ancestry and biobanks. **a)** A scatter plot comparing the ORs across all 106 two-way intersectional contexts for all phenotypes in UKB and AoU European ancestry. **b)** The Spearman’s Rank correlation was calculated between ORs across cohorts for each phenotype, as well as overall. To assess the overall correlation, the mean two-way intersectional OR was subtracted from each cohort’s ORs in order to remove biases arising from differences in PGS predictive power across phenotypes. Bonferroni-corrected significance thresholds were set as * p<.05/3, ** p<.01/3, *** p<.001/3. **c)** Two examples illustrating how ORs change across two variables, their intersections, and cohorts. The left example shows correspondence of ORs for CAD across income and sex between UKB and AoU European ancestry cohorts, while the right example shows non-correspondence for AST across alcohol intake and age. Both examples were statistically significant in the AoU and UKB European ancestry cohorts based on the intersections’ max. OR %Δ. Both the income-by-sex intersection for CAD and the alcohol-by-age intersection for AST were deemed statistically significant (* p<.05/21) in the AoU and UKB European ancestry cohorts, based on the maximum %Δ in ORs at the intersections.

Due to variations in disease prevalence across different contexts, AR estimates for high-PGS individuals within a specific context (*AR*_*c*_) differed substantially from AR estimates for high-PGS individuals overall (*AR*_*unaware*_) (Supplemental Figure 10). While adjusting baseline AR estimates to account for context-specific disease prevalence (*AR*_*adj*_) typically reduces these differences, *AR*_*c*_ can still lead to notable shifts in AR estimates. For high-PGS individuals within European ancestry with low income, *AR*_*c*_ exceeded the unadjusted baseline risk (*AR*_*unaware*_) across all phenotypes, rising by an average of 5.8 percentage-points [95% CI: 5.4, 6.1] in UKB and 3.5 percentage-points [95% CI: 2.8, 4.2] in AoU (Figure 5a). However, when compared to the prevalence-adjusted baseline risk *AR*_*adj*_, their *AR*_*c*_ estimates were lower, decreasing by an average of 1.0 percentage-points [95% CI: −1.3, −.7] in UKB and 1.6 percentage-points [95% CI: −2.3, −.9] in AoU. For MDD, when individuals were further restricted to those additionally with low alcohol intake, the change in estimated AR (*AR*_*c*_-*AR*_*adj*_) was −3.1 percentage-points [95% CI: −4.8, −1.2] and −3.6 percentage-points [95% CI: −6.1, −1.1] in UKB and AoU respectively (Figure 5b).

**Figure 5:**
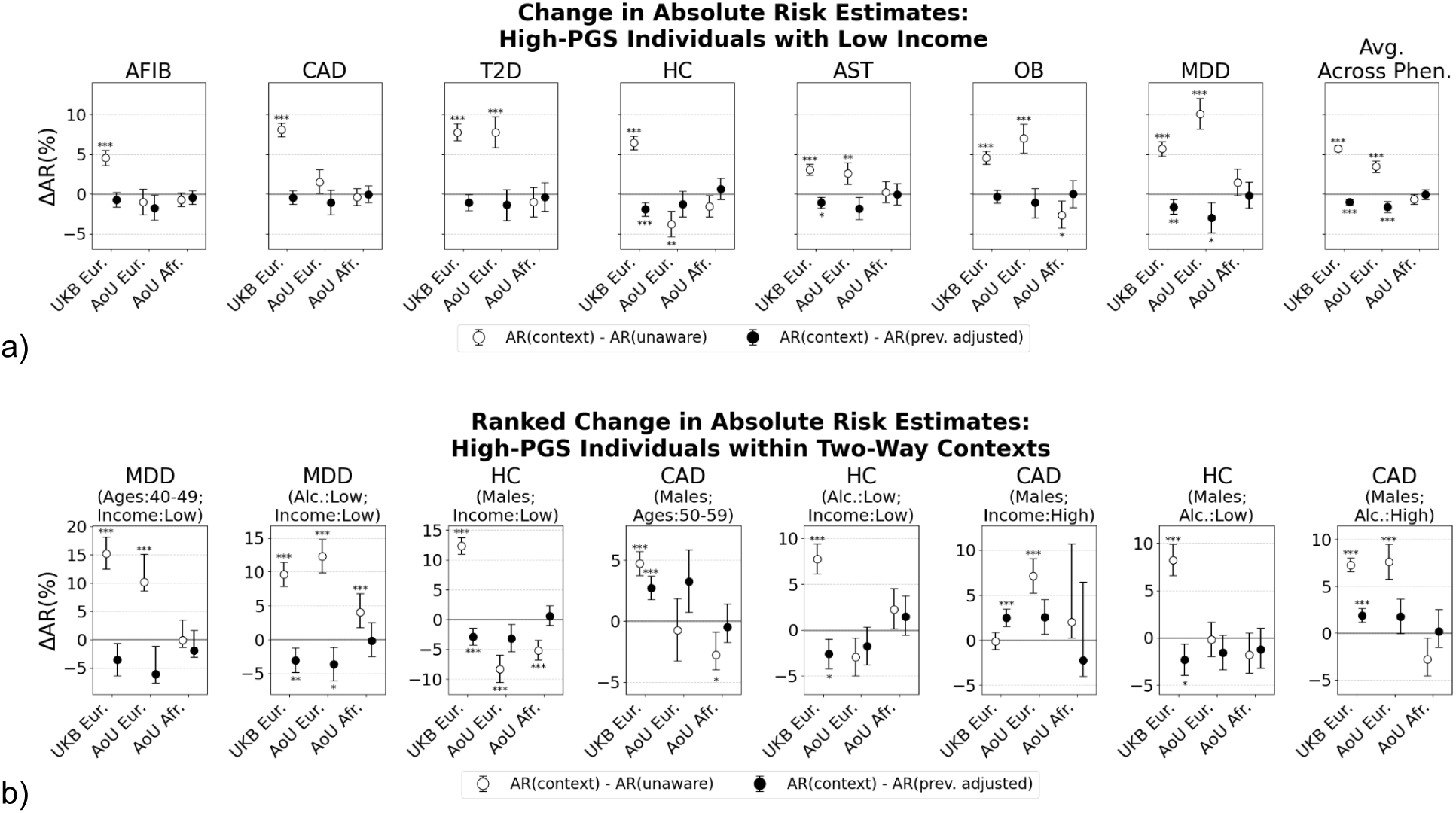
Comparisons of absolute risk estimates with varying levels of context awareness in low-income and intersectional contexts. **a)** A comparison of context-aware absolute risk estimates (*AR*_*c*_) with two baseline risk estimates for high-PGS, low income individuals. The baseline comparisons include the context unaware risk estimate *AR*_*unaware*_ and the prevalence-adjusted baseline risk estimate (context-unaware PGS) *AR*_*adj*_. Results are shown across all phenotypes and as cohort averages for UKB European ancestry, AoU European ancestry, and AoU African ancestry. Statistical significance was determined empirically from a bootstrapped distribution of risk estimates (n=10,000; * p<.05/6, ** p<.01/6, *** p<.001/6). **b)** Comparison of *AR*_*c*_ for high-PGS individuals in the two-way context-phenotype pairs that showed the largest differences between *AR*_*c*_ and *AR*_*adj*_ estimates in UKB that are at least nominally significant (p < .1) in both UKB and AoU European ancestry samples. Statistical significance was determined using a Bonferroni correction for the number of statistical tests within each context-phenotype pair, based on an empirical distribution of bootstrapped risk estimates (n=10,000; * p<.05/6, ** p<.01/6, *** p<.001/6).

## Discussion

Prior research has shown that the predictive power of PGS can be significantly impacted when samples are stratified by context. However, individuals exist within multiple, overlapping contexts, and the influence of these intersectional contexts on PGS utility and clinical relevance has been largely unexplored. To address this gap, we conducted a study examining the impact of contexts on lifetime relative and absolute risk of disease among individuals with high PGS, as defined by the eMERGE consortium. Our study yielded several key findings. First, we found that contexts, especially intersectional contexts, have a significant impact on the relative risk of disease attributable to PGS. Moreover, among the sociodemographic variables examined, those that significantly modified relative genetic risk for a given phenotype were largely consistent across biobanks, although the direction and magnitude of effects across strata within each variable could vary. Finally, absolute risk estimates for high-PGS individuals varied considerably across contexts as well, but differed only moderately from risk estimates that were adjusted for context-specific prevalence.

More specifically, we find that intersectional contexts influence relative genetic risk in two key ways. First, as the number of intersecting contexts increases, the variability in the OR for high-PGS individuals also increases. This upward trend was statistically significant for the majority of phenotypes analyzed in the UKB, reinforcing that the observed increase in variability is not merely a statistical artifact. The absence of a significant trend in T2D and MDD, however, suggests that the impact of intersectional factors on relative genetic risk may be phenotype dependent. Second, in some cases, the interaction between two variables exerts a statistically greater influence on the relative genetic risk of high-PGS individuals than either variable independently. For example, smoking history alone significantly modifies the relative risk of only HC and MDD for high-PGS individuals in UKB. However, smoking history when combined with another variable, significantly modifies relative genetic risk for all phenotypes examined. These findings highlight the complex, non-linear interactions among sociodemographic variables and PGS, emphasizing the need to account for these intersectional context effects in future PGS calibration models.

While the effects of contexts on relative genetic risk vary in direction and magnitude across biobanks, the overall importance of key sociodemographic variables remained largely consistent for individuals with similar genetic ancestry. Specifically, two-way intersectional context effects on relative genetic risk were strongly and significantly correlated across cohorts. The vast majority (25 out of 31) of two-way variable intersections that significantly affected relative genetic risk within AoU European ancestry were also significant in the UKB. The lack of replication in AoU African ancestry may stem from limited statistical power or differences in demographic and environmental variable interactions with PGS. However, we find that even when the same variables significantly modify relative genetic risk in both AoU and UKB for a given phenotype, PGS odds ratios within strata defined by these variables can still differ. This suggests that while the inputs into PGS calibration models may be largely transferable, model performance may decline when applied to different clinical settings, making model recalibration necessary.

Absolute risk estimates for high-PGS individuals within specific contexts differed substantially from context-unaware estimates, though these differences became more modest after adjusting for context-specific prevalences. Similar to contextual effects on relative genetic risk, some intersectional contexts exhibited more pronounced deviations in absolute risk estimates once PGS were contextualized. Among high-PGS individuals with low income and low alcohol intake, the AR for MDD was 3.1 percentage-points lower than the prevalence-adjusted baseline (3.6 percentage-points lower in AoU Eur.), while for CAD in high-PGS males aged 50–59, AR was 2.7 percentage-points higher (3.2 in AoU Eur.). These examples show that for certain phenotypes, failing to contextualize PGS can lead to returning absolute risk estimates that are inappropriately low or high.

Although this study highlights the importance of intersecting contexts in estimating genetic risk, our work is still limited in several ways. First, we were notably underpowered to adequately assess the replicability of our findings in participants of African-like ancestry in AoU. Secondly, we imposed relatively arbitrary bins to categorize continuous contextual factors. This results in reduced statistical power, but improved interpretability and visualization of our findings. Finally, in practice, modeling the effects of intersecting contexts is challenging. With limited sample sizes available for calibration model fitting and a large number of cross-context interaction terms, regression models can be constrained by low statistical power, possible non-identifiability, and issues with positivity for binary traits. Especially for disease traits with low prevalence, many intersecting contexts may not contain any cases. Despite these limitations, we are still able to identify statistically significant intersections of contextual factors that impact PGS performance across multiple traits, several of which were replicable in an external matched-ancestry sample.

Notably, this study employed an absolute risk model of disease prevalence rather than disease incidence. This distinction is important in clinical settings, as the results are less suited for prevention or early risk stratification and more relevant to assessing current disease burden. Additionally, contextualizing PGS may have a greater impact on predicting incident risk than observed for prevalent risk. Addressing these and other limitations will require the development of methods that can effectively leverage complex contextual interactions to improve individual-level prediction and calibrate uncertainty around genetic risk estimates. If PGS are to be integrated into clinical practice, accounting for context should ensure greater fairness in the translation of genomic findings towards improving health and well-being for all.

## Data Availability

Access to used datasets require an application through the UK Biobank and the All of Us Researcher Workbench.

https://www.researchallofus.org

http://www.ukbiobank.ac.uk

## Declaration of Interests

JWS is a member of the Scientific Advisory Board of Sensorium Therapeutics (with options), has received consulting fees from Data Driven, Inc., and Tempus, Inc., and has received grant support from Biogen, Inc.

## Acknowledgments

This research has been conducted using the UK Biobank Resource under Application Number 32568. The All of Us Research Program is supported by the National Institutes of Health, Office of the Director: Regional Medical Centers: 1 OT2 OD026549; 1 OT2 OD026554; 1 OT2 OD026557; 1 OT2 OD026556; 1 OT2 OD026550; 1 OT2 OD 026552; 1 OT2 OD026553; 1 OT2 OD026548; 1 OT2 OD026551; 1 OT2 OD026555; IAA #: AOD 16037; Federally Qualified Health Centers: HHSN 263201600085U; Data and Research Center: 5 U2C OD023196; Biobank: 1 U24 OD023121; The Participant Center: U24 OD023176; Participant Technology Systems Center: 1 U24 OD023163; Communications and Engagement: 3 OT2 OD023205; 3 OT2 OD023206; and Community Partners: 1 OT2 OD025277; 3 OT2 OD025315; 1 OT2 OD025337; 1 OT2 OD025276. In addition, the All of Us Research Program would not be possible without the partnership of its participants. JWS is supported in part by U01HG008685 and R01MH137218. TG is supported in part by R01HG12354. MC and JDT were supported by the Mass General Brigham Training Program in Precision and Genomic Medicine (T32HG010464).

## Data and code availability

This study uses individual-level genotype and phenotype data from the UK Biobank and the All of Us Research Program’s Controlled Tier Dataset V7. Access to these datasets require an application through the UK Biobank (http://www.ukbiobank.ac.uk) and the All of Us Researcher Workbench (https://www.researchallofus.org). An exception to the Data and Statistics Dissemination Policy was granted by the All of Us Resource Access Board for the results reported in this manuscript.

Software used to perform the main analysis in this manuscript are available via Github at: https://github.com/MihaelCudic/PGS_in_contexts.

## Supplement

**Supplemental Table 1a:** Participant counts and case numbers for each phenotype studied in UK Biobank. Only participants of European-British-like ancestry were included to control for potential effects of population stratification.

| Phenotype | Abbreviation | Condition Code | PGS Weights | Participants | # of Cases |
| --- | --- | --- | --- | --- | --- |
| Atrial Fibrillation | AFIB | I48 (ICD-10) | AFIB: Nielsen et al., 2018 <sup>21</sup> | 375,054 | 16,921 |
| Coronary Artery Disease | CAD | I21-I25 (ICD-10), K40-K46, K49, K50, K75, (OPCS-4) or self-reported illness code 1075 | CAD: Nikpay et al., 2015 <sup>22</sup> | 375,054 | 28,840 |
| Type 2 Diabetes | T2D | E11 (ICD-10) or self-reported illness code 1223 | T2D: Scott et al., 2017 <sup>23</sup> | 375,054 | 20,761 |
| Hypercholesterolemia | HC | E78.0 (ICD-10) | LDL Cholesterol: Willer et al., 2013 <sup>23</sup> | 375,054 | 35,810 |
| Asthma | AST | J45 (ICD-10) | AST: Demenais et al., 2017 <sup>24</sup> | 375,054 | 28,772 |
| Obesity | OB | E66 (ICD-10) | BMI: Locke et al., 2015 <sup>25</sup> | 375,054 | 20,707 |
| Major Depressive Disorder | MDD | F32, F33, F34, F38, F38 (ICD-10), or self-reported illness code 1286 | MDD: Wray et al., 2018 (excluding UKB) <sup>25</sup> | 375,054 | 31,754 |

**Supplemental Table 1b:** Participant counts and case numbers for each phenotype studied in All of Us.

| Phenotype | Condition Code (SNOMED) | PGS Weights | African-like Ancestry |  | European-like Ancestry |  |
| --- | --- | --- | --- | --- | --- | --- |
|  |  |  | Participants | # of Cases | Participants | # of Cases |
| Atrial Fibrillation | 313217 | AFIB: Nielsen et al., 2018 <sup>21</sup> | 36,552 | 1,484 | 99,477 | 9,193 |
| Coronary Artery Disease | 317576 | CAD: Nikpay et al., 2015 <sup>22</sup> | 36,552 | 3,614 | 99,477 | 15,501 |
| Type 2 Diabetes | 201826 | T2D: Scott et al., 2017 <sup>23</sup> | 36,552 | 8,346 | 99,477 | 16,253 |
| Hypercholesterolemia | 4029305 | LDL Cholesterol: Willer et al., 2013 <sup>23</sup> | 36,552 | 4,354 | 99,477 | 20,924 |
| Asthma | 317009 | AST: Demenais et al., 2017 <sup>24</sup> | 36,552 | 5,485 | 99,477 | 14,389 |
| Obesity | 433736 | BMI: Locke et al., 2015 <sup>25</sup> | 36,552 | 9,451 | 99,477 | 23,847 |
| Major Depressive Disorder | 4152280 | MDD: Wray et al., 2018 (excluding UKB) <sup>25</sup> | 36,552 | 6,137 | 99,477 | 20,453 |

**Supplemental Table 2:** Synchronized environmental and case numbers across AoU and UKB cohorts.

|  | Dataset: | All of Us |  |  |  |  | UK Biobank |  |  |  |
| --- | --- | --- | --- | --- | --- | --- | --- | --- | --- | --- |
| Environ. Categories | Sync. Labels | Q. Code | Q. Abbrev. | Answer | # of Samples (Afr.) | # of Samples (Eur.) | Data Field | Q. Abbrev. | Answer | # of Samples (Eur.) |
| Sex | Females |  | Sex | Female | 19,384 | 55,940 | 31 | Sex | Female | 202,280 |
|  | Males |  |  | Male | 16,154 | 41,333 |  |  | Male | 172,774 |
| Age | 40-49 |  | Age | 40-49 | 7,834 | 13,372 | 21022 | Age | 40-49 | 84,265 |
|  | 50-59 |  |  | 50-59 | 12,761 | 20,236 |  |  | 50-59 | 124,510 |
|  | 60-69 |  |  | 60+ | 9,543 | 28,387 |  |  | 60+ | 166,277 |
| Monthly Alcohol Intake | Low | 1586201: How often did you have a drink containing alcohol in the past year? | Drink Freq. Past Year | ≤1 | 16,303 | 43,725 | 1558: About how often do you drink alcohol? | Monthly Alcohol Intake | ≤ 1 | 64,625 |
|  | Med. |  |  | 1 to 8 | 5,722 | 18,274 |  |  | 1 to 8 | 139,652 |
|  | High |  |  | >8 | 6,172 | 31,806 |  |  | ≥12 | 170,267 |
| Smoking History | Low | 1585857: Have you smoked at least 100 cigarettes in your entire life? (There are 20 cigarettes in a pack.)? | 100 Cigs. Lifetime | Yes | 16,470 | 53,022 | 20116 | Smoking Status | No History | 201,738 |
|  | High |  |  | No | 18,614 | 44,177 |  |  | Previous & Current | 171,745 |
| Income | Low | 1585375: What is your annual household income from all sources? | Ann. Income | <\$35k | 19,740 | 20,338 | 738: What is the average total income before tax received by your household? | House. Income | ≤£18 | 70,175 |
| | Med. | | | \$35k to \$100k | 5,045 | 32,314 | | | £18-£52k | 167,090 |
| | High | | | >\$100k | 1,590 | 32,266 | | | ≥£52k | 85,652 |
| Deprivation | Low | Deprivation Index derived from American Community Survey, linked by 3-digit ZIP code | Neigh. Deprivation. | ≤0.3 | 5,649 | 52,896 | 22189 | Townsend Deprivation. | ≤-3 | 141,325 |
|  | Med. |  |  | -0.3 to 0.375 | 13,126 | 34,332 |  |  | -3 to 0 | 133,498 |
|  | High |  |  | ≥0.375 | 17,752 | 12,202 |  |  | ≥0 | 99,784 |

**Supplemental Table 3:** PGS cutoff points for all 7 phenotypes as proposed by eMERGE. Note that a recommended cutoff for MDD was not given, so we used 1% to include the phenotype in our study.

| Phenotype | eMERGE Cutoff<br>Top % |
| --- | --- |
| Atrial Fibrillation | 3% |
| Coronary Artery Disease | 5% |
| Type 2 Diabetes | 2% |
| Hypercholesterolemia | 3% |
| Asthma | 5% |
| Obesity | 3% |
| Major Depressive Disorder<sup>†</sup> | 1% |

**Supplemental Figure 1:**
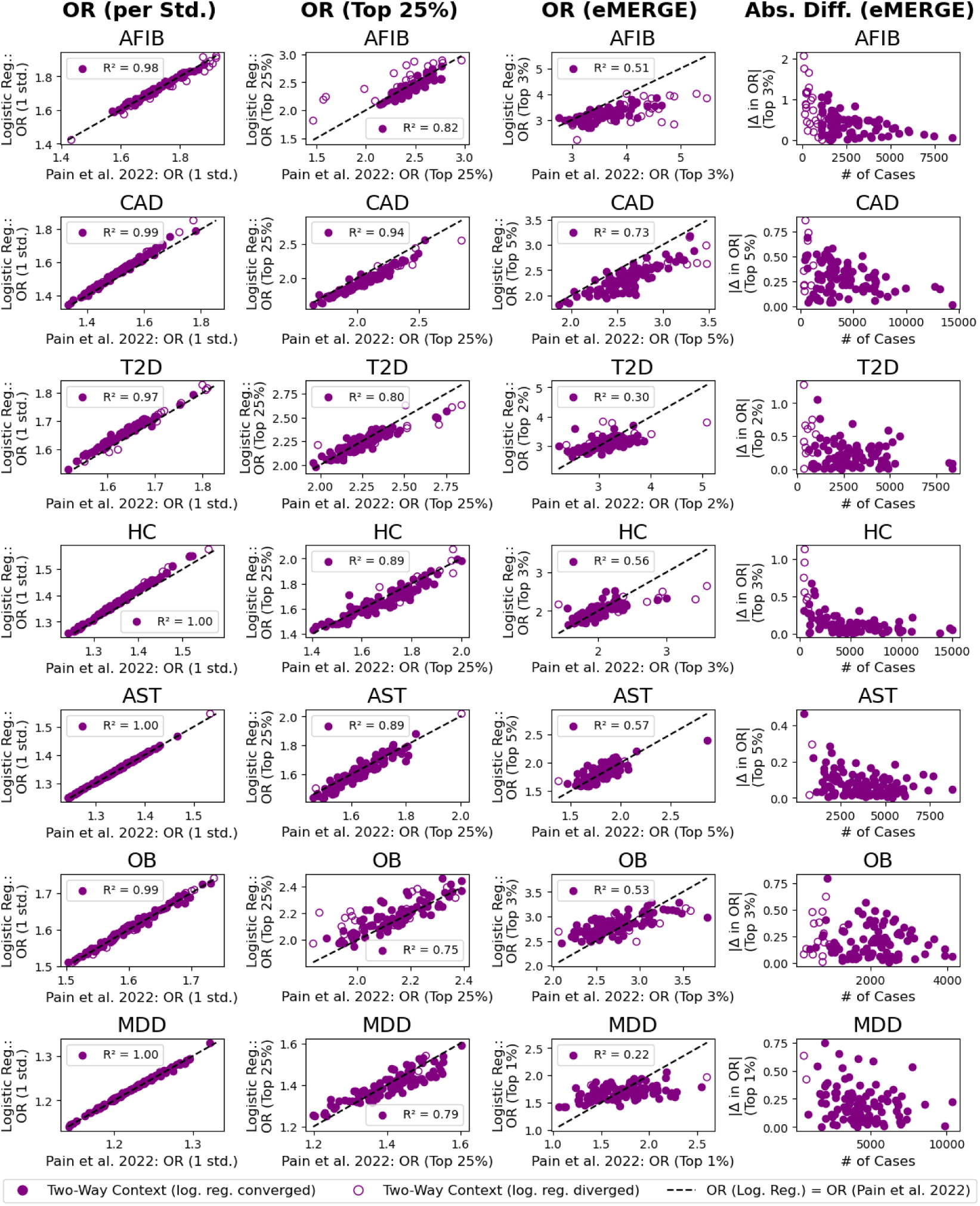
Comparisons of all 106 two-way intersectional ORs in UKB at varying cutoffs using two different methods. The first method fits a logistic regression model from the PGS, covariates, and principal components (PCs) as predictors. The second method, which is used throughout the main study, first regresses out covariates and PCs from both the phenotype and the PGS using the full sample. It then converts the resulting effect sizes to ORs using the method described by Pain et al. (2022). The first three columns show ORs per standard deviation of PGS, for the top 25% of PGS, and for the top eMERGE-defined PGS percentile, respectively. The final column shows the absolute difference in ORs from the high-PGS group (third column) as a function of case count. Open circles (○) indicate non-converged logistic models due to low sample size. R² values are shown for converged points in the first three columns.

**Supplemental Figure 2:**
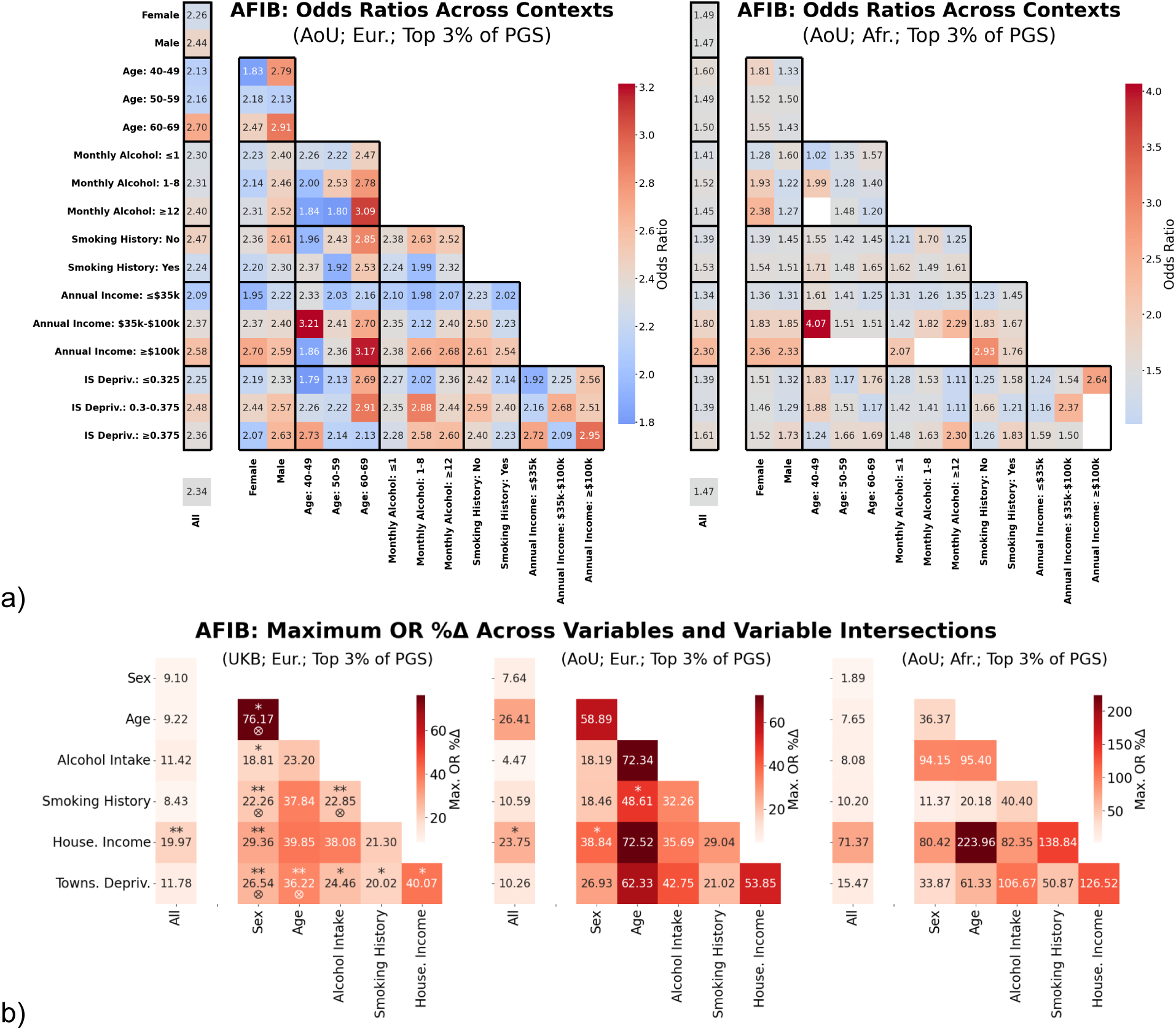
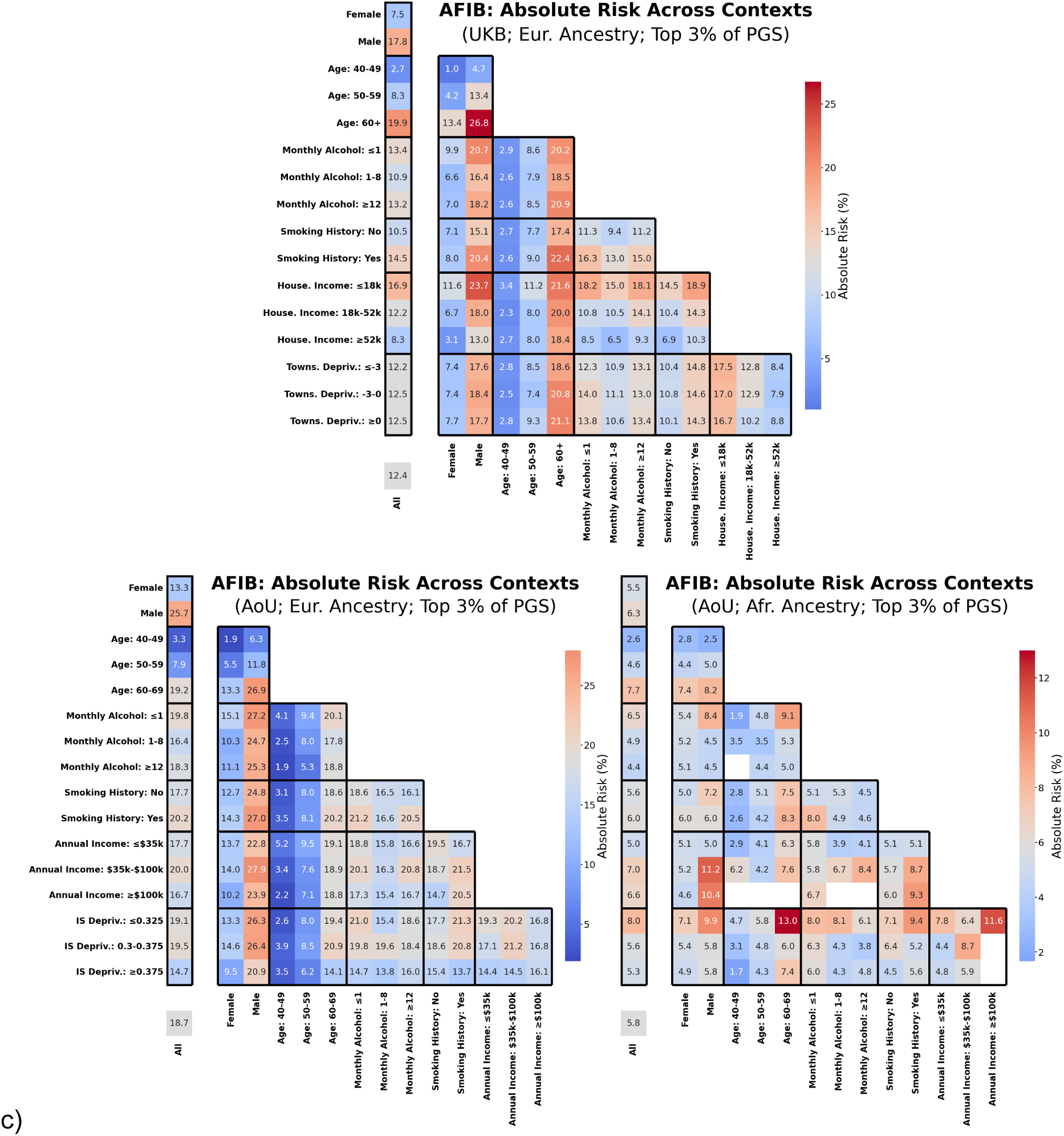
Lifetime relative and absolute risk of atrial fibrillation in high-PGS individuals across contexts. The lifetime odds ratios **(a)** and absolute risks **(c)** of atrial fibrillation (AFIB) for individuals in the top 3% of the PGS distribution, as determined by the recommended eMERGE PGS cutoff, across varying primary and two-way contexts for three cohorts: UKB European ancestry, AoU European ancestry, and AoU African ancestry. Relative risk estimates for AFIB within the UK Biobank (UKB) European ancestry cohort are shown in the main manuscript (Figure 2a). Missing cells indicate fewer than 20 cases, in which case the analysis was skipped due to insufficient statistical power. Odds ratio and absolute risk was calculated using Pain et al. (2022) approximation. **b)** The maximum percent difference in odds ratio (max. OR %Δ) observed across strata, calculated either within a single variable (i.e., across all strata associated with that variable) or at the intersection of two variables (i.e., across all strata formed by combinations of two variables), for all three cohorts.

**Supplemental Figure 3:**
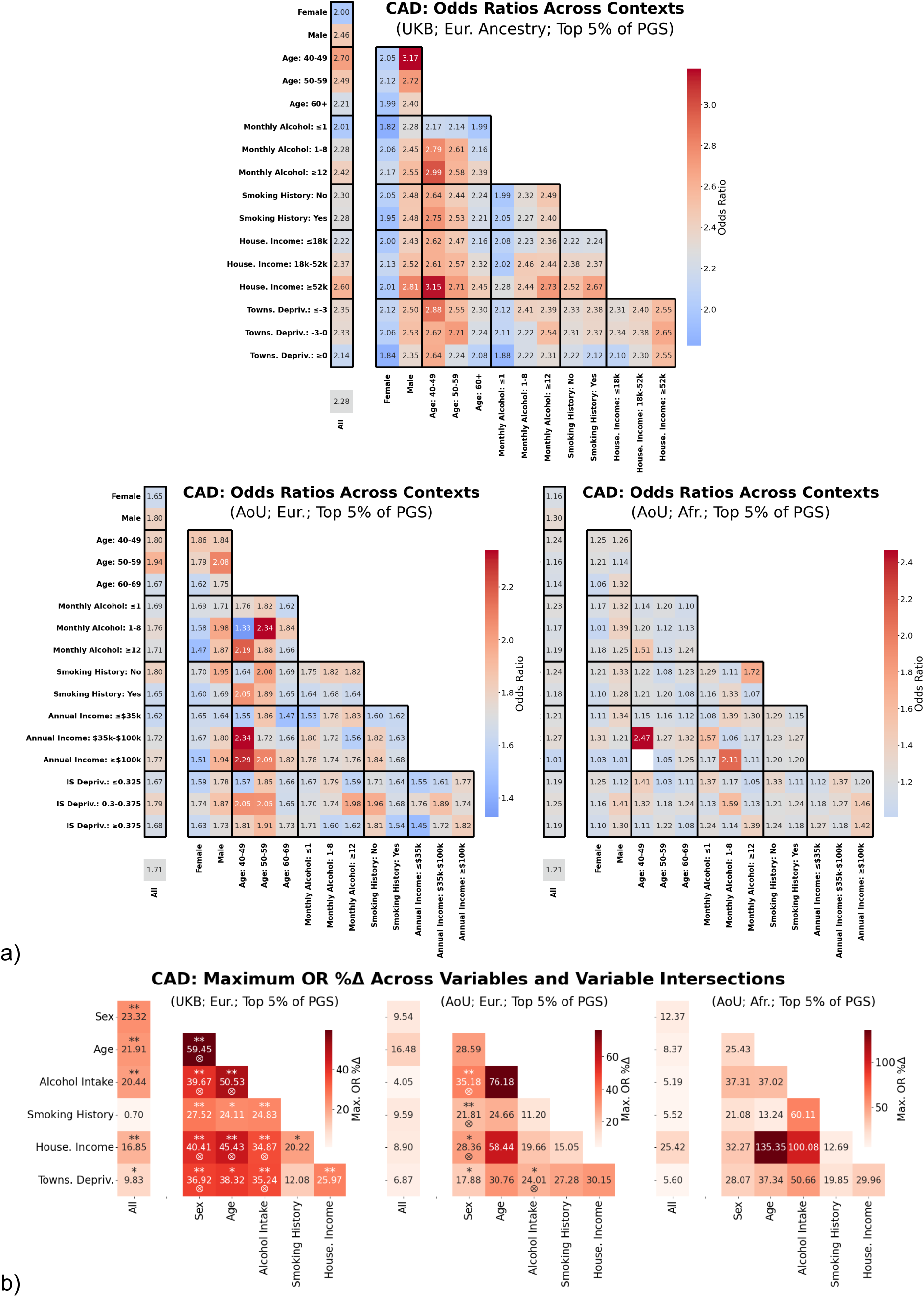
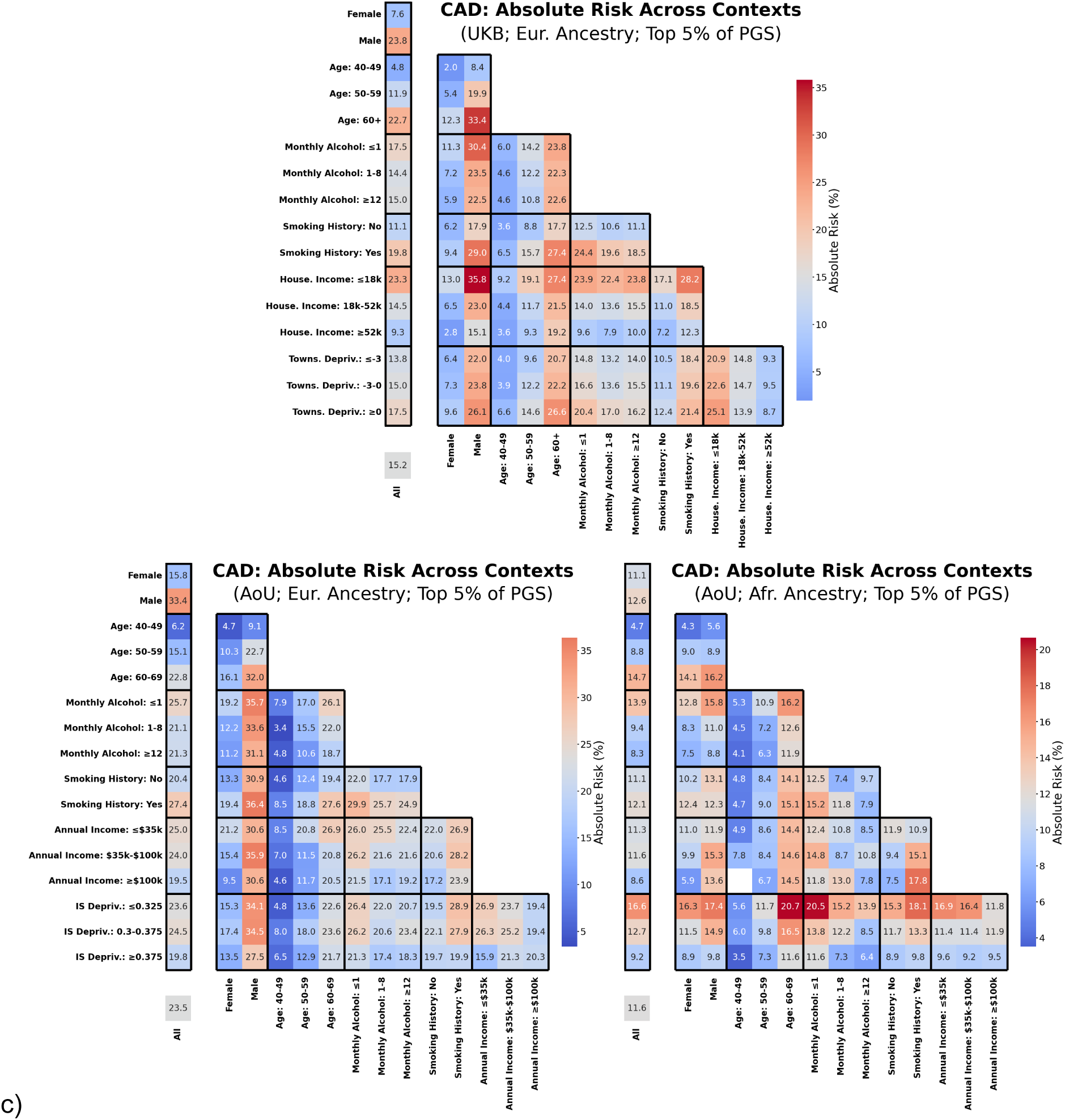
Lifetime relative and absolute risk of coronary artery disease in high-PGS individuals across contexts. The lifetime odds ratios **(a)** and absolute risks **(c)** of coronary artery disease (CAD) for individuals in the top 5% of the PGS distribution, as determined by the recommended eMERGE PGS cutoff, across varying primary and two-way contexts for three cohorts: UKB European ancestry, AoU European ancestry, and AoU African ancestry. Missing cells indicate fewer than 20 cases, in which case the analysis was skipped due to insufficient statistical power. Odds ratio and absolute risk was calculated using Pain et al. (2022) approximation. **b)** The maximum percent difference in odds ratio (max. OR %Δ) observed across strata, calculated either within a single variable (i.e., across all strata associated with that variable) or at the intersection of two variables (i.e., across all strata formed by combinations of two variables), for all three cohorts.

**Supplemental Figure 4:**
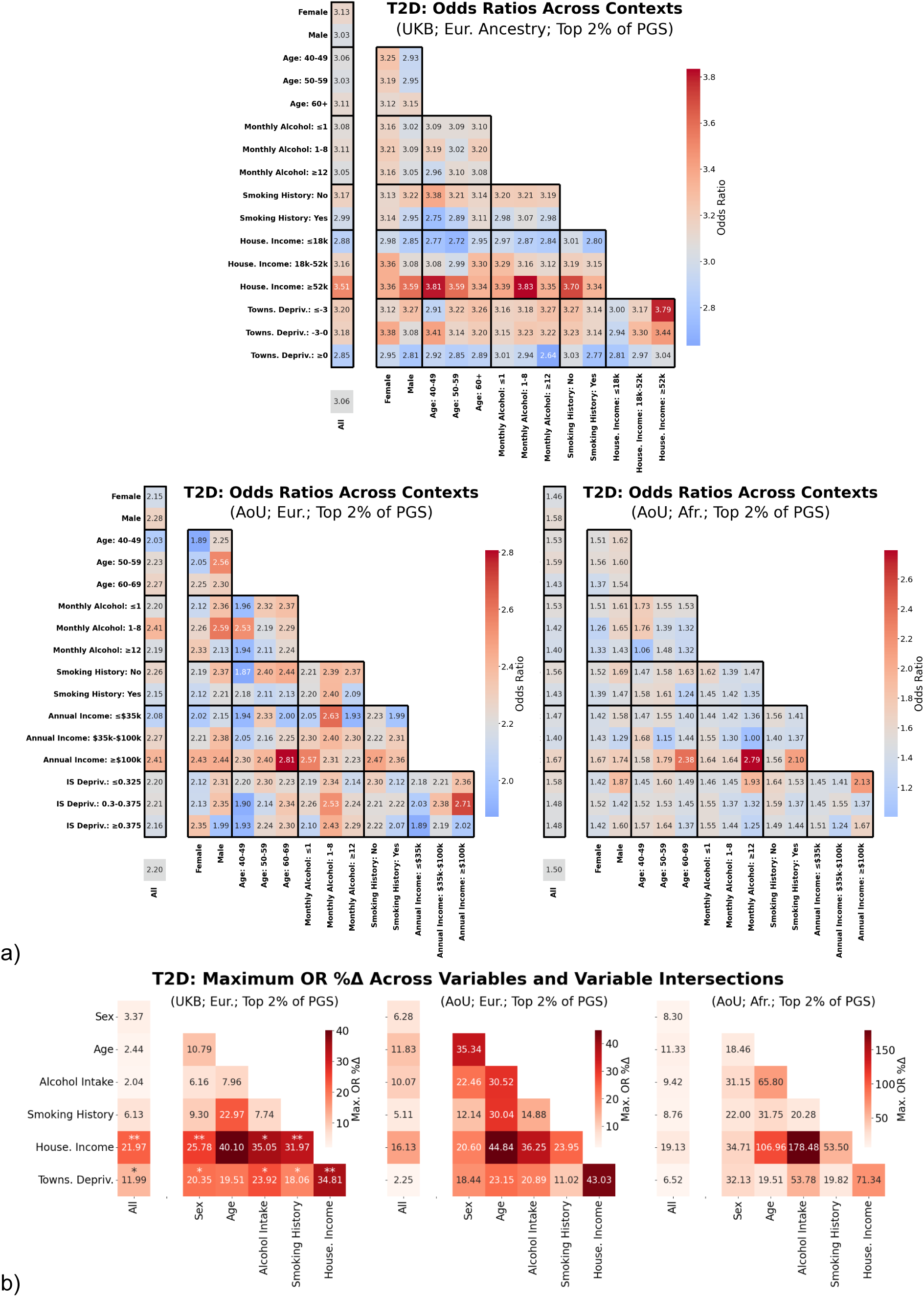
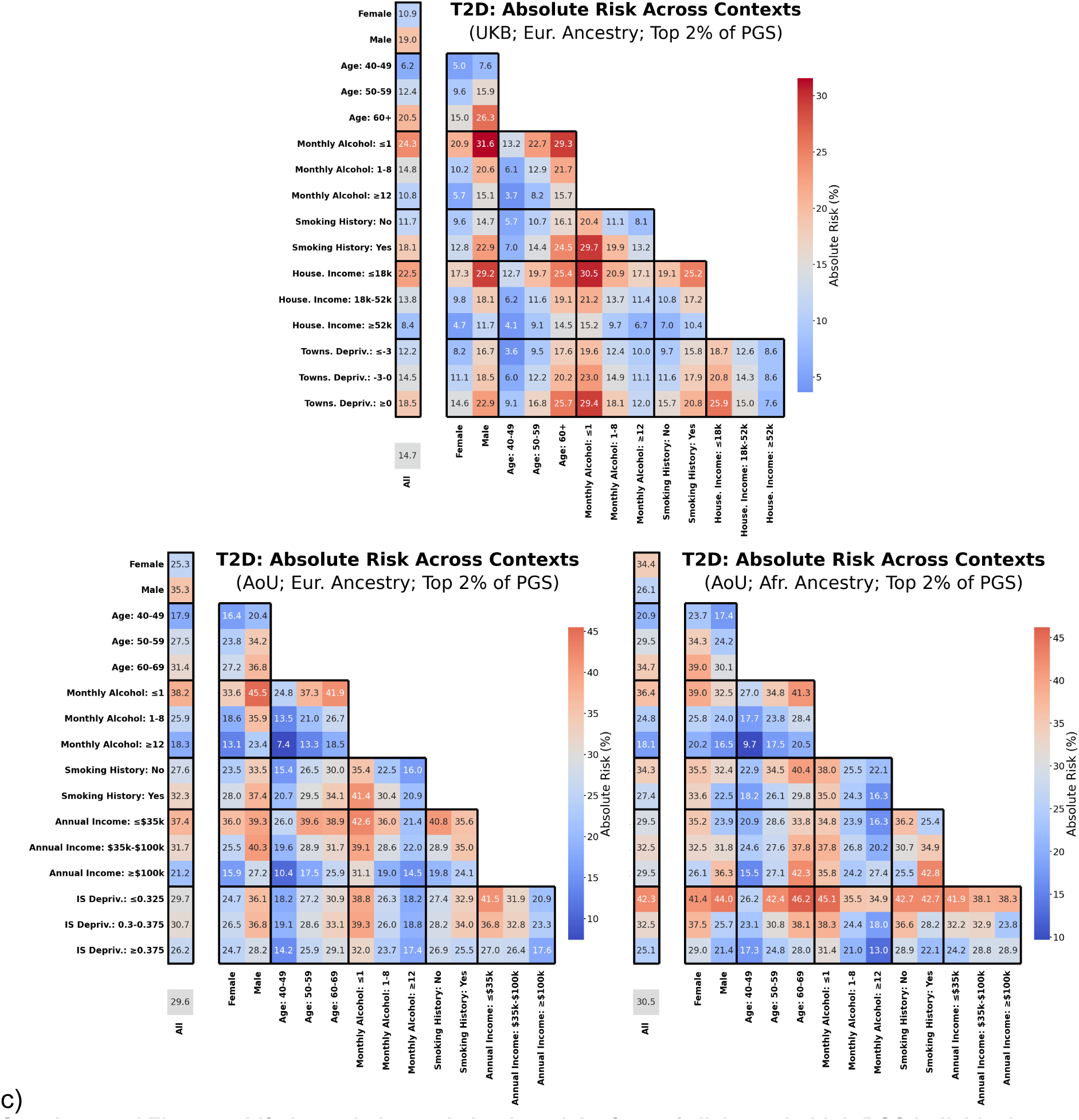
Lifetime relative and absolute risk of type 2 diabetes in high-PGS individuals across contexts. The lifetime odds ratios **(a)** and absolute risks **(c)** of type 2 diabetes (T2D) for individuals in the top 2% of the PGS distribution, as determined by the recommended eMERGE PGS cutoff, across varying primary and two-way contexts for three cohorts: UKB European ancestry, AoU European ancestry, and AoU African ancestry. Odds ratio and absolute risk was calculated using Pain et al. (2022) approximation. **b)** The maximum percent difference in odds ratio (max. OR %Δ) observed across strata, calculated either within a single variable (i.e., across all strata associated with that variable) or at the intersection of two variables (i.e., across all strata formed by combinations of two variables), for all three cohorts.

**Supplemental Figure 5:**
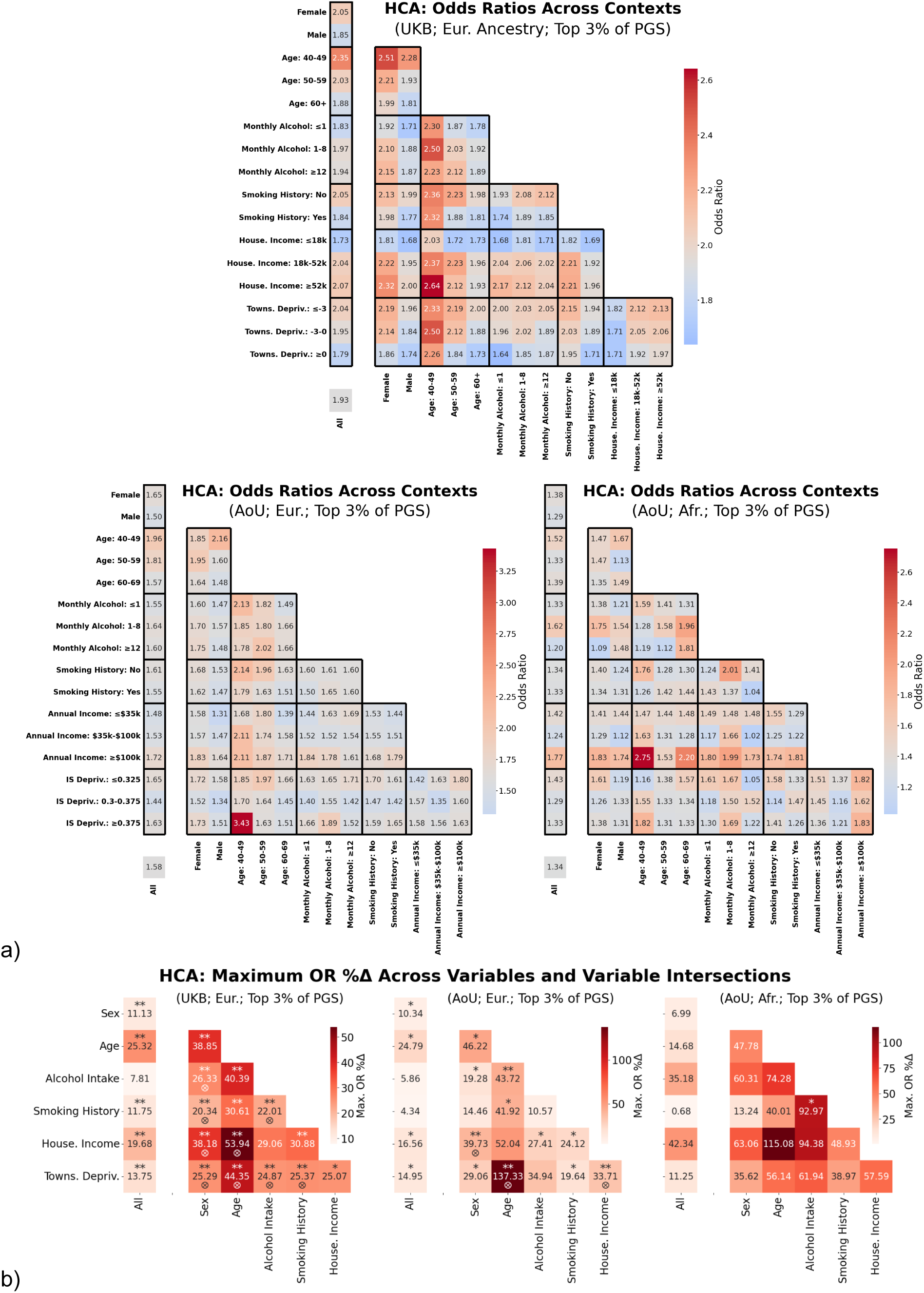
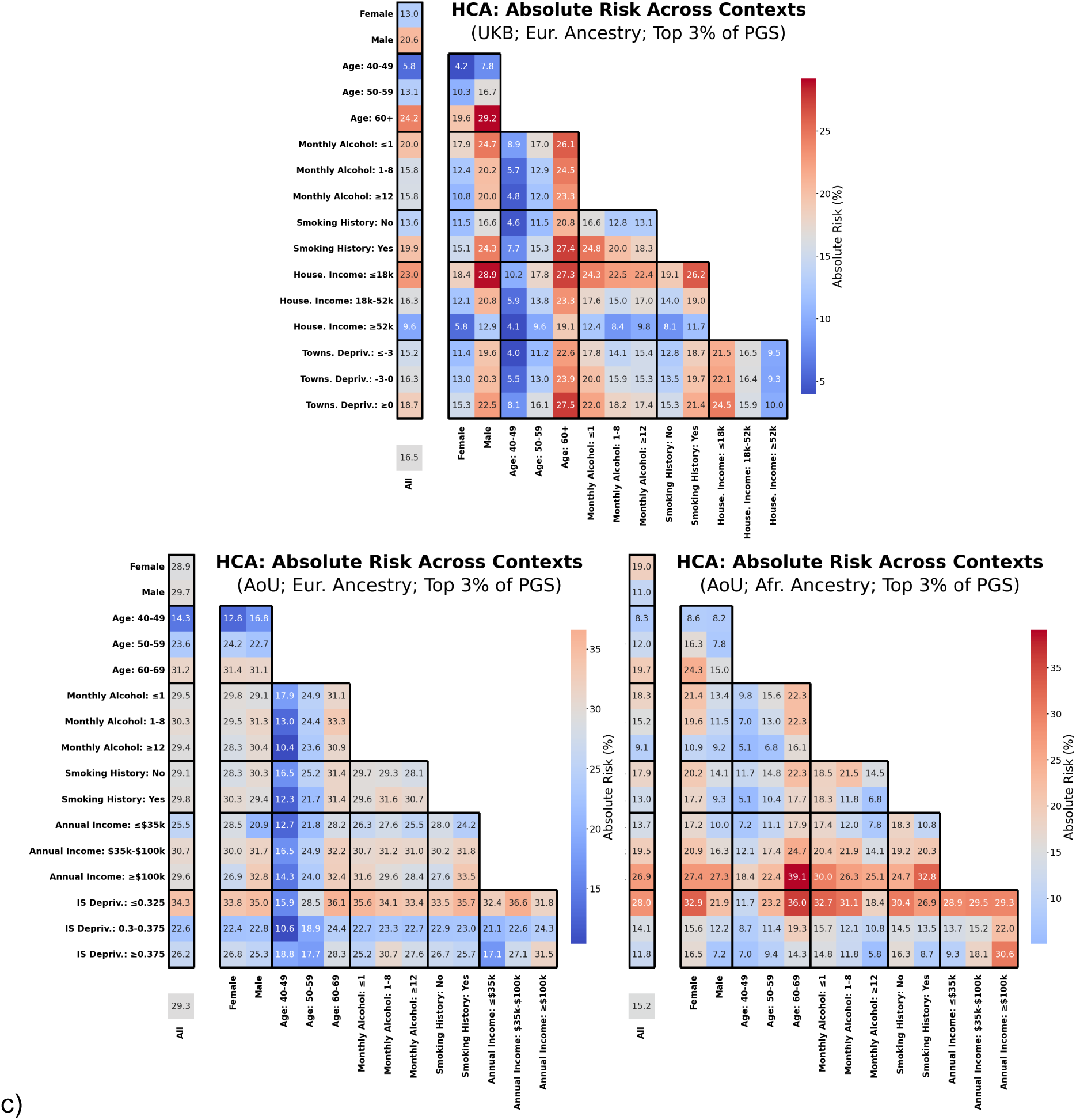
Lifetime relative and absolute risk of hypercholesterolemia in high-PGS individuals across contexts. The lifetime odds ratios **(a)** and absolute risks **(c)** of hypercholesterolemia (HC) for individuals in the top 3% of the PGS distribution, as determined by the recommended eMERGE PGS cutoff, across varying primary and two-way contexts for three cohorts: UKB European ancestry, AoU European ancestry, and AoU African ancestry. Odds ratio and absolute risk was calculated using Pain et al. (2022) approximation. **b)** The maximum percent difference in odds ratio (max. OR %Δ) observed across strata, calculated either within a single variable (i.e., across all strata associated with that variable) or at the intersection of two variables (i.e., across all strata formed by combinations of two variables), for all three cohorts.

**Supplemental Figure 6:**
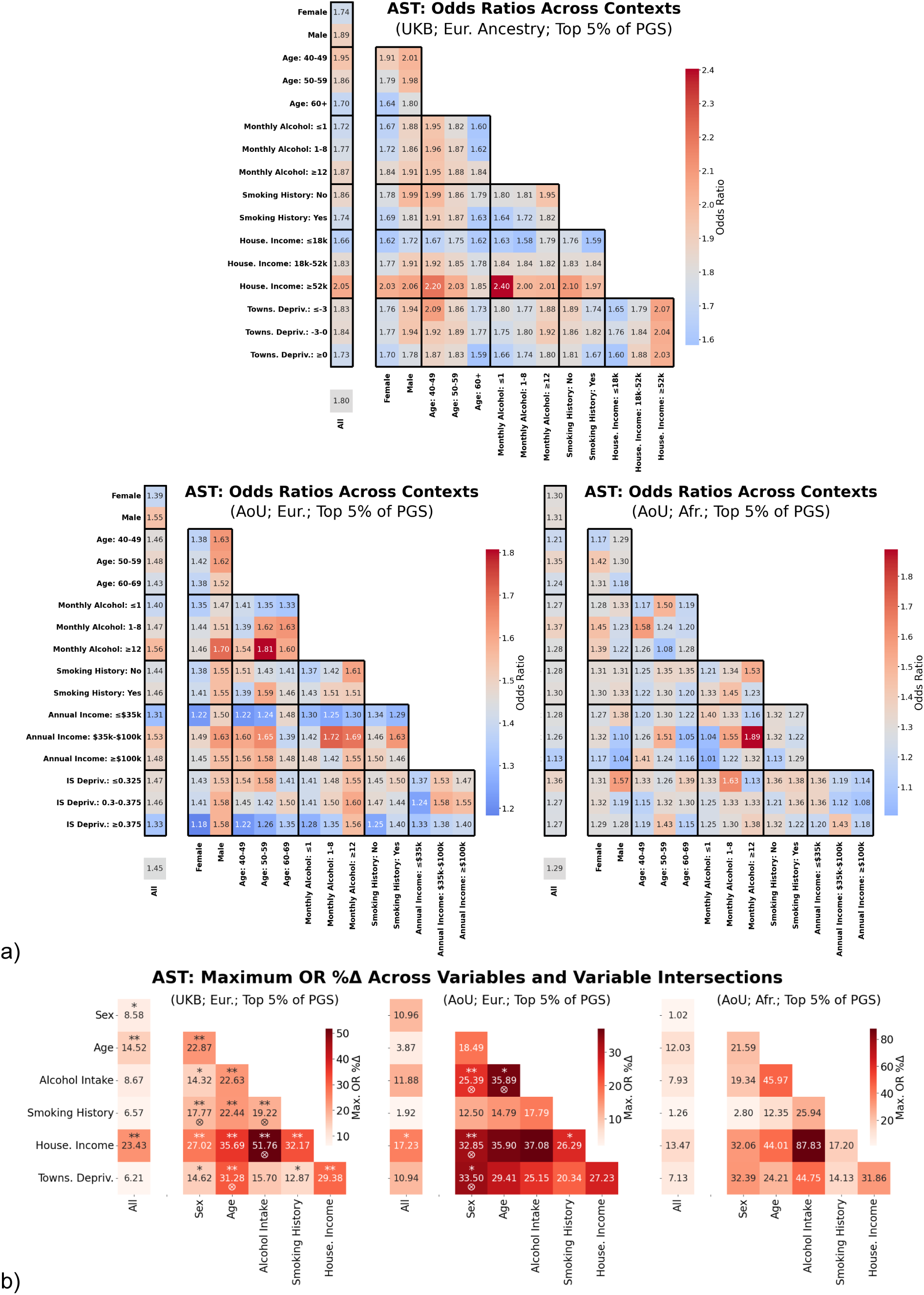
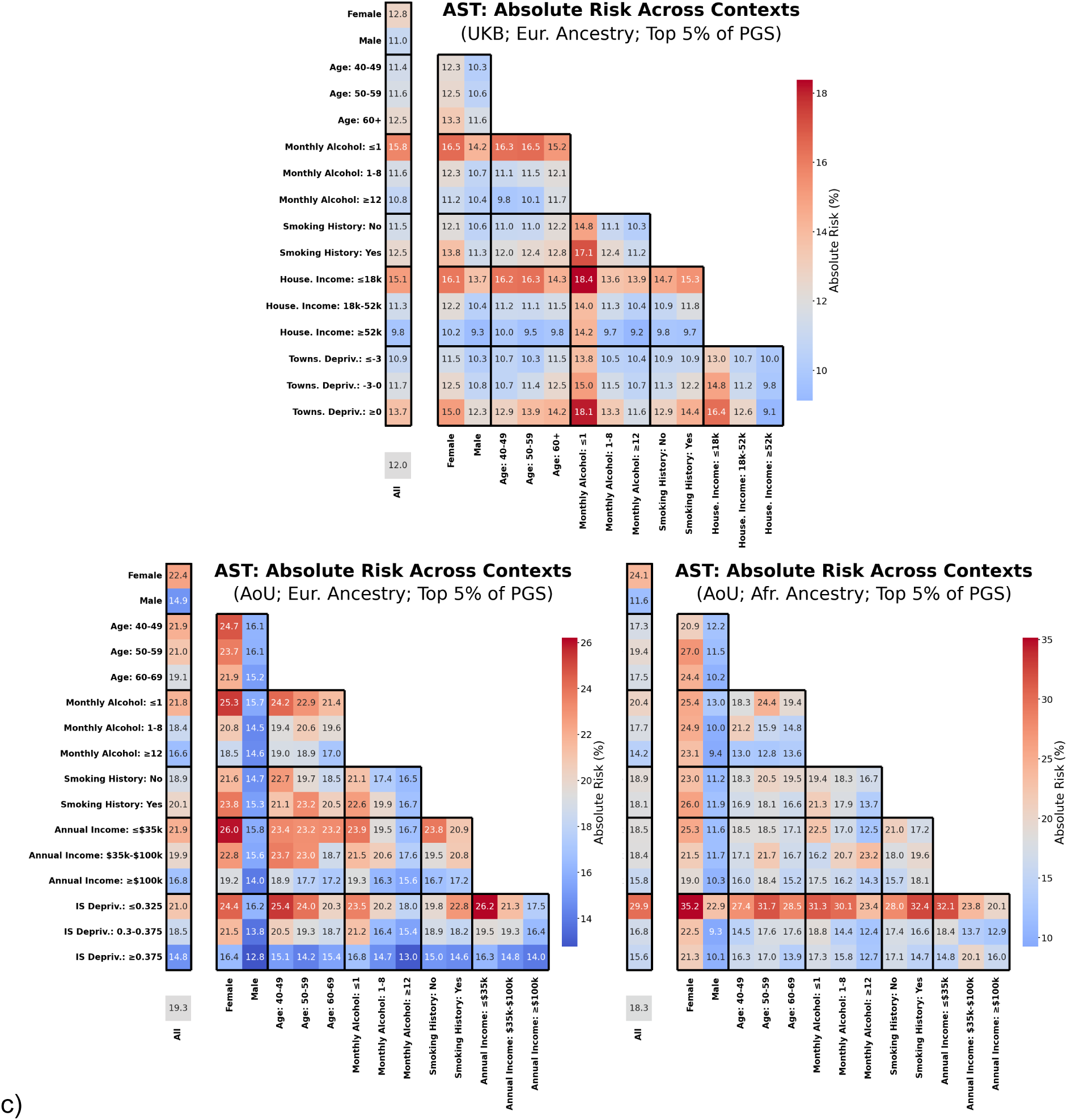
Lifetime relative and absolute risk of asthma in high-PGS individuals across contexts. The lifetime odds ratios **(a)** and absolute risks **(c)** of asthma (AST) for individuals in the top 5% of the PGS distribution, as determined by the recommended eMERGE PGS cutoff, across varying primary and two-way contexts for three cohorts: UKB European ancestry, AoU European ancestry, and AoU African ancestry. Odds ratio and absolute risk was calculated using Pain et al. (2022) approximation. **b)** The maximum percent difference in odds ratio (max. OR %Δ) observed across strata, calculated either within a single variable (i.e., across all strata associated with that variable) or at the intersection of two variables (i.e., across all strata formed by combinations of two variables), for all three cohorts.

**Supplemental Figure 7:**
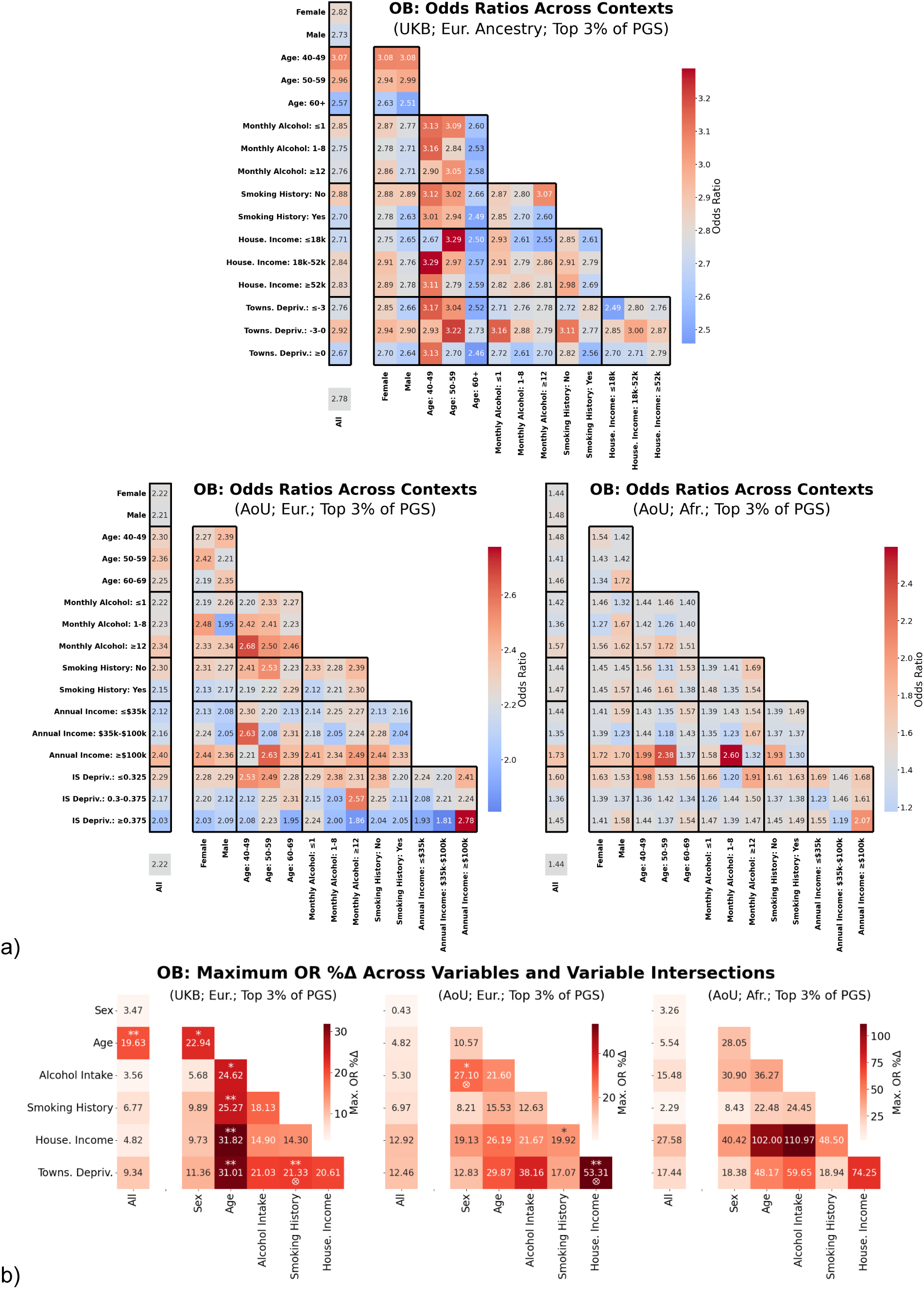
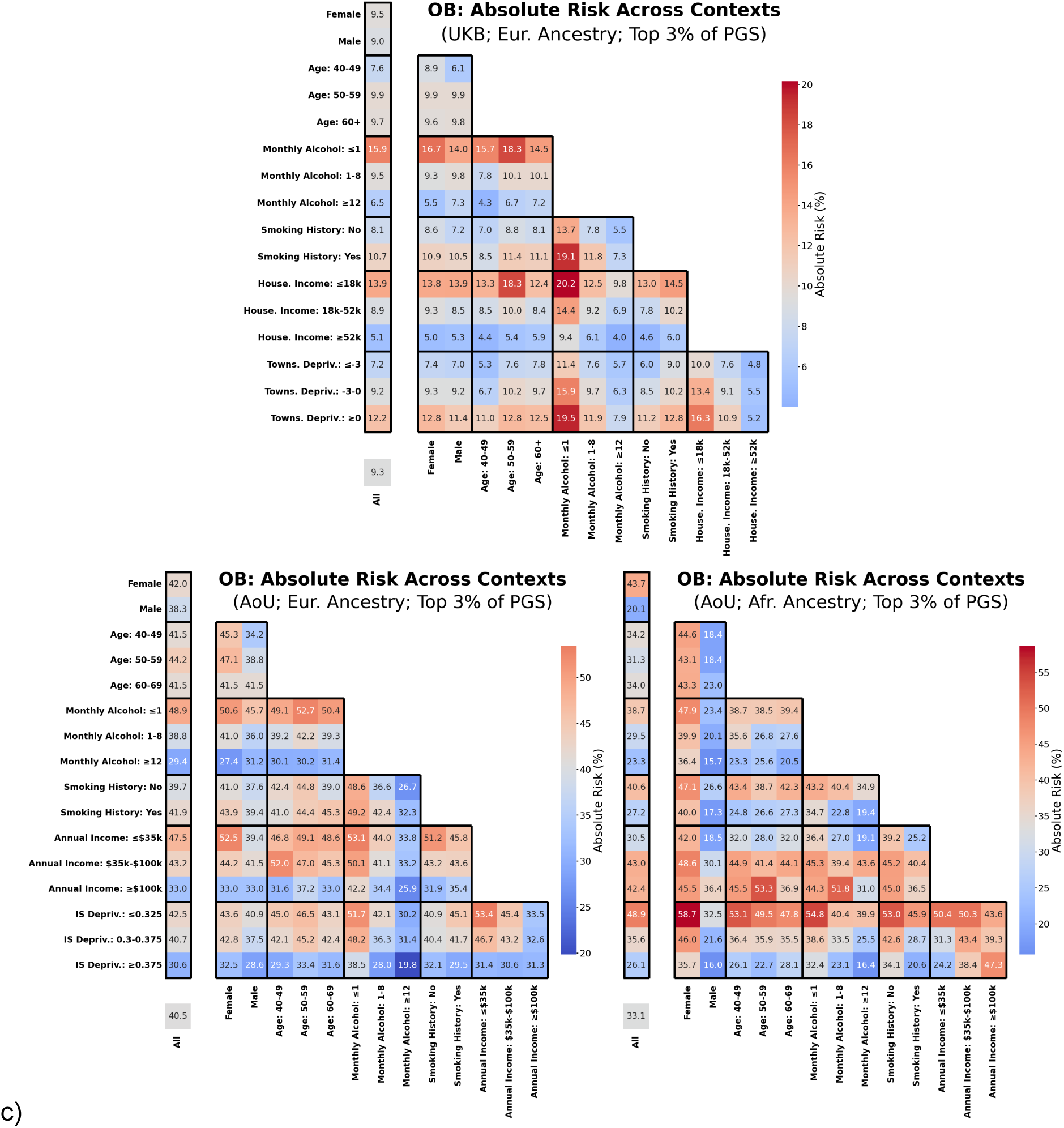
Lifetime relative and absolute risk of obesity in high-PGS individuals across contexts. The lifetime odds ratios **(a)** and absolute risks **(c)** of obesity (OB) for individuals in the top 2% of the PGS distribution, as determined by the recommended eMERGE PGS cutoff, across varying primary and two-way contexts for three cohorts: UKB European ancestry, AoU European ancestry, and AoU African ancestry. Odds ratio and absolute risk was calculated using Pain et al. (2022) approximation. **b)** The maximum percent difference in odds ratio (max. OR %Δ) observed across strata, calculated either within a single variable (i.e., across all strata associated with that variable) or at the intersection of two variables (i.e., across all strata formed by combinations of two variables), for all three cohorts.

**Supplemental Figure 8:**
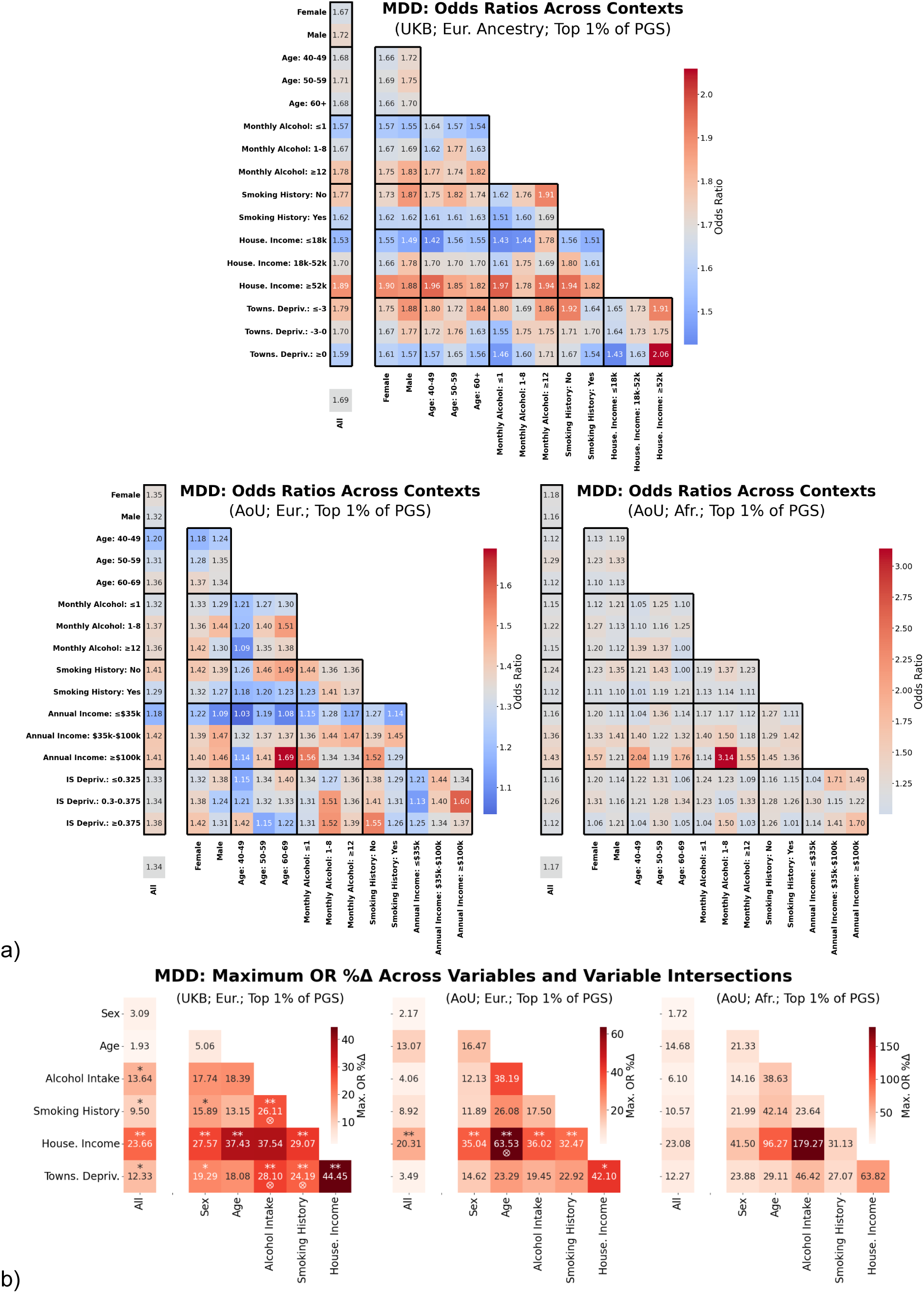
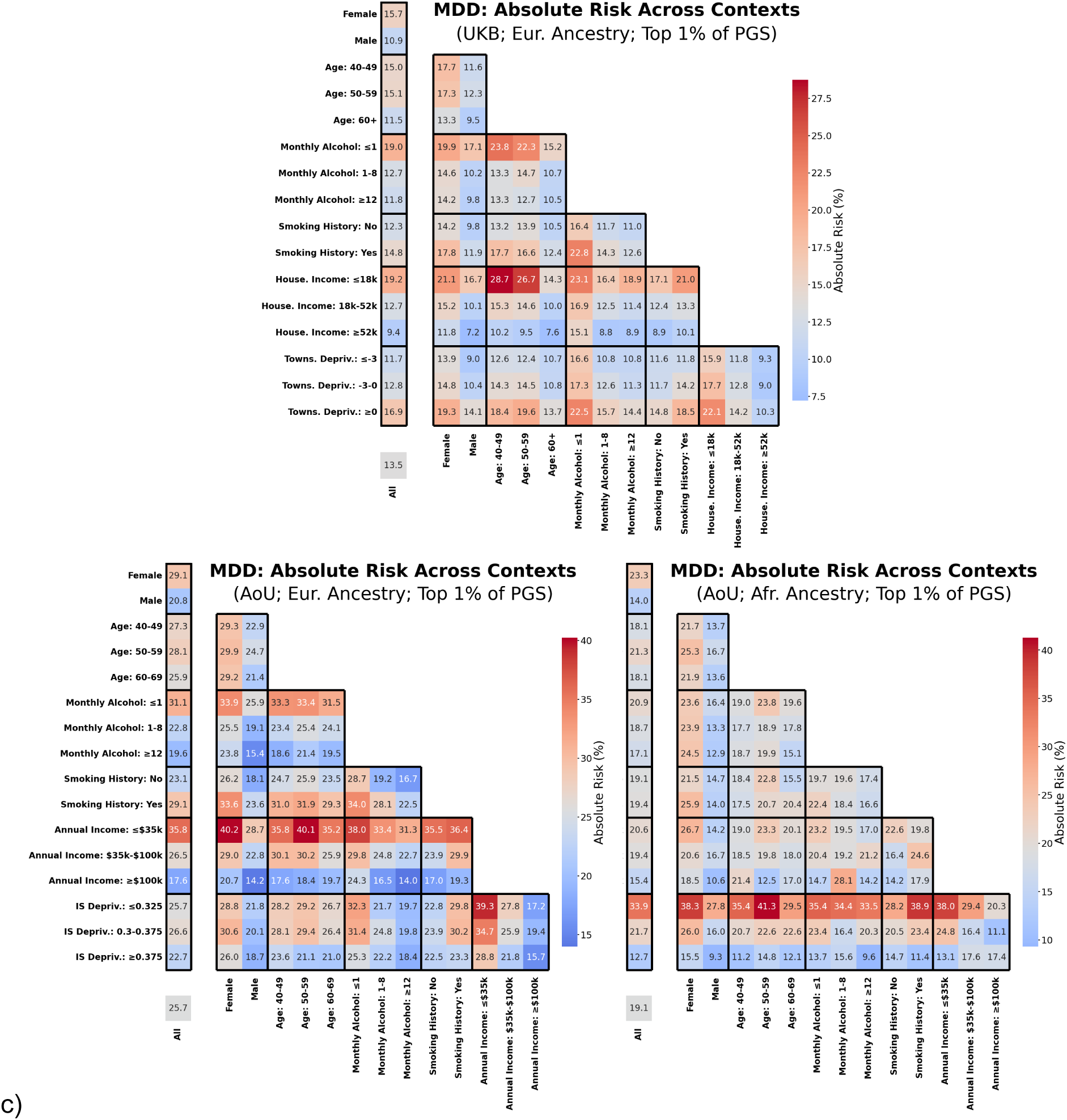
Lifetime relative and absolute risk of major depression in high-PGS individuals across contexts. The lifetime odds ratios **(a)** and absolute risks **(c)** of major depressive disorder (MDD) for individuals in the top 1% of the PGS distribution across varying primary and two-way contexts for three cohorts: UKB European ancestry, AoU European ancestry, and AoU African ancestry. Unlike other phenotypes in this study, eMERGE did not recommend a PGS cutoff for MDD, so a 1% cutoff was used instead. Odds ratio and absolute risk was calculated using Pain et al. (2022) approximation. **b)** The maximum percent difference in odds ratio (max. OR %Δ) observed across strata, calculated either within a single variable (i.e., across all strata associated with that variable) or at the intersection of two variables (i.e., across all strata formed by combinations of two variables), for all three cohorts.

**Supplemental Figure 9:**
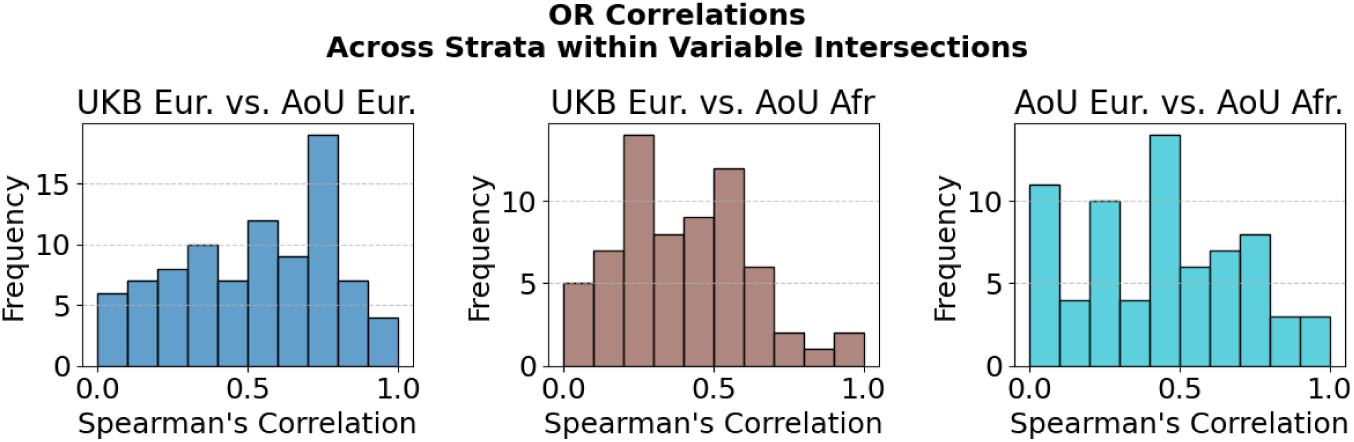
Distributions of the OR Spreaman’s Rank correlations between cohorts for strata within each two-way variable intersections. Each distribution contains 105 correlations in total, representing all 15 possible two-way variable intersections across the 7 phenotypes.

**Supplemental Figure 10:**
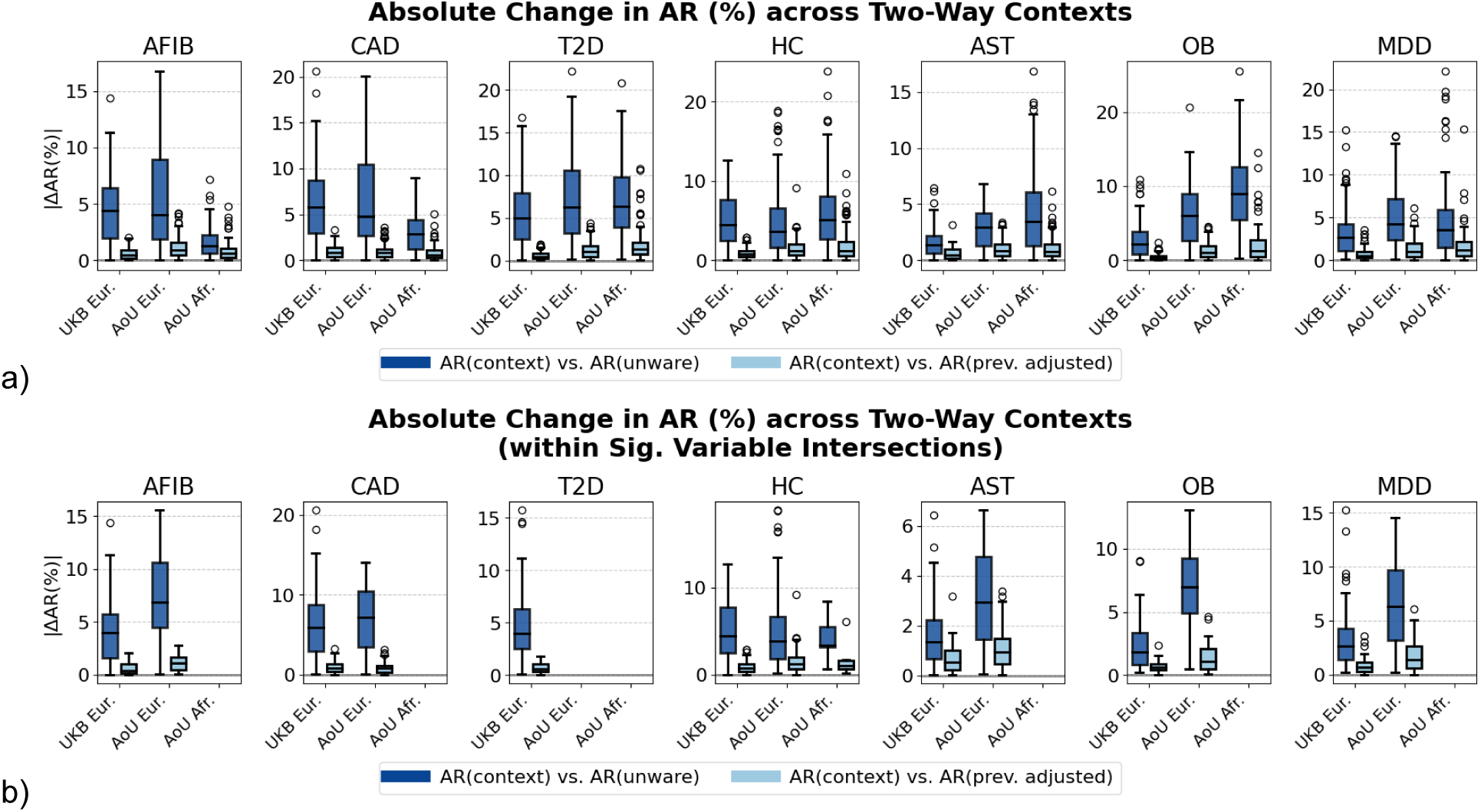
Distribution of absolute risk estimate differences across two-way intersectional contexts. a) The distribution of absolute differences in context-aware risk estimates *AR*_*c*_ from two baseline risk estimates, across all 106 two-way contexts. The baseline comparisons include the context-unaware risk estimate *AR*_*unaware*_ and the prevalence adjusted risk estimate *AR*_*adj*_. b) Same as (a), but restricted to the subset of two-way contexts that lie within variable intersections identified as significant for each cohort in the main analysis.

## Supplementary Files

**Supplemental_Table_S1.xlsx**: A comprehensive table containing odds ratios (OR), absolute risks (both *AR*_*c*_ and prevalence adjusted baseline *AR*_*adj*._), condition prevalence, and total sample size for each context and for each phenotype. Data are stratified across all one-way, two-way, and three-way contexts and are provided separately for each cohort. Note that three-way intersecting contexts were not analyzed in AoU due to smaller sample sizes. Additionally, contexts in AoU with 20 or fewer individuals with a condition were excluded (i.e., represented as blank cells) in accordance with AoU’s dissemination policy.

**Supplemental_Table_S2.xlsx**: A summary table presenting the maximum percentage change in odds ratios (ORs) for each phenotype across individual variables and their intersections. The table includes 95% confidence intervals from bootstrapped distributions (n = 10,000), p-values comparing the estimates to zero, and p-values comparing each estimate to the corresponding point estimate in its parent contexts (p-value Interx.).

## Notes

### Author Declarations

This study uses individual-level genotype and phenotype data from the UK Biobank and the All of Us Research Program's Controlled Tier Dataset V7. An exception to the Data and Statistics Dissemination Policy was granted by the All of Us Resource Access Board for the results reported in this manuscript.

## Bibliography

1. Lewis, C.M., and Vassos, E. (2020). Polygenic risk scores: From research tools to clinical instruments. Genome Med. 12, 1–11.

2. O’Sullivan, J.W., Raghavan, S., Marquez-Luna, C., Luzum, J.A., Damrauer, S.M., Ashley, E.A., O’Donnell, C.J., Willer, C.J., Natarajan, P., and American Heart Association Council on Genomic and Precision Medicine; Council on Clinical Cardiology; Council on Arteriosclerosis, Thrombosis and Vascular Biology; Council on Cardiovascular Radiology and Intervention; Council on Lifestyle and Cardiometabolic Health; and Council on Peripheral Vascular Disease (2022). Polygenic Risk Scores for Cardiovascular Disease: A Scientific Statement From the American Heart Association. Circulation 146, e93–e118.

3. Vassy, J.L., Brunette, C.A., Lebo, M.S., MacIsaac, K., Yi, T., Danowski, M.E., Alexander, N.V.J., Cardellino, M.P., Christensen, K.D., Gala, M., et al. (2023). The GenoVA study: Equitable implementation of a pragmatic randomized trial of polygenic-risk scoring in primary care. Am J Hum Genet 110, 1841–1852.

4. Hao, L., Kraft, P., Berriz, G.F., Hynes, E.D., Koch, C., Korategere V, Kumar, P., Parpattedar, S.S., Steeves, M., Yu, W., Antwi, A.A., et al. (2022). Development of a clinical polygenic risk score assay and reporting workflow. Nat Med 28, 1006–1013.

5. Khera, A.V., Chaffin, M., Aragam, K.G., Haas, M.E., Roselli, C., Choi, S.H., Natarajan, P., Lander, E.S., Lubitz, S.A., Ellinor, P.T., et al. (2018). Genome-wide polygenic scores for common diseases identify individuals with risk equivalent to monogenic mutations. Nat. Genet. 50, 1219–1224.

6. Lennon, N.J., Kottyan, L.C., Kachulis, C., Abul-Husn, N.S., Arias, J., Belbin, G., Below, J.E., Berndt, S.I., Chung, W.K., Cimino, J.J., et al. (2024). Selection, optimization and validation of ten chronic disease polygenic risk scores for clinical implementation in diverse US populations. Nat. Med. 30, 480–487.

7. Wang, Y., Kanai, M., Tan, T., Kamariza, M., Tsuo, K., Yuan, K., Zhou, W., Okada, Y., Huang, H., Turley, P., et al. (2023). Polygenic prediction across populations is influenced by ancestry, genetic architecture, and methodology. Cell Genom 3, 100408.

8. Kachuri, L., Chatterjee, N., Hirbo, J., Schaid, D.J., Martin, I., Kullo, I.J., Kenny, E.E., Pasaniuc, B., Witte, J.S., and Ge, T. (2024). Principles and methods for transferring polygenic risk scores across global populations. Nat. Rev. Genet. 25, 8–25.

9. Martin, A.R., Kanai, M., Kamatani, Y., Okada, Y., Neale, B.M., and Daly, M.J. (2019). Clinical use of current polygenic risk scores may exacerbate health disparities. Nat. Genet. 51, 584–591.

10. Wang, Y., Guo, J., Ni, G., Yang, J., Visscher, P.M., and Yengo, L. (2020). Theoretical and empirical quantification of the accuracy of polygenic scores in ancestry divergent populations. Nat. Commun. 11, 3865.

11. Hou, K., Xu, Z., Ding, Y., Harpak, A., and Pasaniuc, B. (2023). Calibrated prediction intervals for polygenic scores across diverse contexts. medRxiv.

12. Ding, Y., Hou, K., Xu, Z., Pimplaskar, A., Petter, E., Boulier, K., Privé, F., Vilhjálmsson, B.J., Olde Loohuis, L.M., and Pasaniuc, B. (2023). Polygenic scoring accuracy varies across the genetic ancestry continuum. Nature 1–8.

13. Hui, D., Dudek, S., Kiryluk, K., Walunas, T.L., Kullo, I.J., Wei, W.-Q., Tiwari, H.K., Peterson, J.F., Chung, W.K., Davis, B., et al. (2024). Risk factors affecting polygenic score performance across diverse cohorts. medRxiv.

14. Mostafavi, H., Harpak, A., Conley, D., Pritchard, J.K., and Przeworski, M. (2019). Variable prediction accuracy of polygenic scores within an ancestry group. bioRxiv 9, 629949.

15. Bycroft, C., Freeman, C., Petkova, D., Band, G., Elliott, L.T., Sharp, K., Motyer, A., Vukcevic, D., Delaneau, O., O’Connell, J., et al. (2018). The UK Biobank resource with deep phenotyping and genomic data. Nature 562, 203–209.

16. Ge, T., Chen, C.-Y., Ni, Y., Feng, Y.-C.A., and Smoller, J.W. (2019). Polygenic prediction via Bayesian regression and continuous shrinkage priors. Nat. Commun. 10, 1776.

17. Ge, T., Irvin, M.R., Patki, A., Srinivasasainagendra, V., Lin, Y.-F., Tiwari, H.K., Armstrong, N.D., Benoit, B., Chen, C.-Y., Choi, K.W., et al. (2022). Development and validation of a trans-ancestry polygenic risk score for type 2 diabetes in diverse populations. Genome Med. 14, 70.

18. Chang, C.C., Chow, C.C., Tellier, L.C., Vattikuti, S., Purcell, S.M., and Lee, J.J. (2015). Second-generation PLINK: rising to the challenge of larger and richer datasets. Gigascience 4, 7.

19. All of Us Research Program Genomics Investigators (2024). Genomic data in the All of Us Research Program. Nature 627, pages 340–346.

20. Pain, O., Gillett, A.C., Austin, J.C., Folkersen, L., and Lewis, C.M. (2022). A tool for translating polygenic scores onto the absolute scale using summary statistics. Eur. J. Hum. Genet. 1–10.

21. Nielsen, J.B., Thorolfsdottir, R.B., Fritsche, L.G., Zhou, W., Skov, M.W., Graham, S.E., Herron, T.J., McCarthy, S., Schmidt, E.M., Sveinbjornsson, G., et al. (2018). Biobank-driven genomic discovery yields new insight into atrial fibrillation biology. Nat. Genet. 50, 1234–1239.

22. the CARDIoGRAMplusC4D Consortium (2015). A comprehensive 1000 Genomes–based genome-wide association meta-analysis of coronary artery disease. Nat. Genet. 47, 1121–1130.

23. Scott, R.A., Scott, L.J., Mägi, R., Marullo, L., Gaulton, K.J., Kaakinen, M., Pervjakova, N., Pers, T.H., Johnson, A.D., Eicher, J.D., et al. (2017). An expanded genome-wide association study of type 2 diabetes in Europeans. Diabetes 66, 2888–2902.

24. Demenais, F., Margaritte-Jeannin, P., Barnes, K.C., Cookson, W.O.C., Altmüller, J., Ang, W., Barr, R.G., Beaty, T.H., Becker, A.B., Beilby, J., et al. (2018). Multiancestry association study identifies new asthma risk loci that colocalize with immune-cell enhancer marks. Nat. Genet. 50, 42–53.

25. Locke, A.E., The LifeLines Cohort Study, Kahali, B., Berndt, S.I., Justice, A.E., Pers, T.H., Day, F.R., Powell, C., Vedantam, S., Buchkovich, M.L., et al. (2015). Genetic studies of body mass index yield new insights for obesity biology. Nature 518, 197–206.

